# Clinical stratification of Major Depressive Disorder in the UK Biobank: A gene-environment-brain Topological Data Analysis

**DOI:** 10.1101/2024.09.19.24313867

**Authors:** Emma Tassi, Alessandro Pigoni, Nunzio Turtulici, Federica Colombo, Lidia Fortaner-Uyà, Anna Maria Bianchi, Francesco Benedetti, Chiara Fabbri, Benedetta Vai, Paolo Brambilla, Eleonora Maggioni

**Author notes:** Paolo Brambilla, Department of Neurosciences and Mental Health, Fondazione IRCCS Ca’ Granda Ospedale Maggiore Policlinico, Milan, Italy., **Email:**.

## Abstract

Major depressive disorder (MDD) is a leading cause of disability worldwide, affecting over 300 million people and posing a significant burden on healthcare systems. MDD is highly heterogeneous, with variations in symptoms, treatment response, and comorbidities that could be determined by diverse etiologic mechanisms, including genetic and neural substrates, and societal factors.

Characterizing MDD subtypes with distinct clinical manifestations could improve patient care through targeted personalized interventions. Recently, Topological Data Analysis (TDA) has emerged as a promising tool for identifying homogeneous subgroups of diverse medical conditions and key disease markers, reducing complex data into comprehensible representations and capturing essential dataset features.

Our study applied TDA to data from the UK Biobank MDD subcohort composed of 3052 samples, leveraging genetic, environmental, and neuroimaging data to stratify MDD into clinically meaningful subtypes. TDA graphs were built from unimodal and multimodal feature sets and quantitatively compared based on their capability to predict depression severity, physical comorbidities, and treatment response outcomes.

Our findings showed a key role of the environment in determining the severity of depressive symptoms. Comorbid medical conditions of MDD were best predicted by brain imaging characteristics, while brain functional patterns resulted the best predictors of treatment response profiles.

Our results suggest that considering genetic, environmental, and brain characteristics is essential to characterize the heterogeneity of MDD, providing avenues for the definition of robust markers of health outcomes in MDD.

## 1. Introduction

Major depressive disorder (MDD) is one of the most prevalent mental illnesses and among the leading causes of disability worldwide, affecting over 300 million people and representing a major burden in health systems ^1–3^. MDD is a highly heterogeneous disorder, not only phenotypically (e.g., symptoms and response to treatment), but also in terms of etiologic mechanisms, such as genetic, and neural substrates, societal and environmental factors ^4,5^.

The incomplete understanding of the determinants of this heterogeneity, especially in biological terms, limits drug discovery and treatment personalization for patients with MDD^6,7^. Regarding treatment, no single drug for depression is universally effective and sequential antidepressant trials are often needed, with about one third of patients not showing sufficient symptoms relief after the first trial, and about 15% of patients still symptomatic even after multiple trials^7,8^. This trial-and-error approach increases the time needed to reach remission, therefore greatly increasing the overall burden of the disorder^9^. Lack of response to at least two antidepressants with adequate duration and dose is defined as treatment-resistant depression (TRD), a condition associated with significant disability and socioeconomic burden ^10,11^. The identification and characterization of homogeneous subtypes of MDD, with distinct patterns of clinical manifestations and outcome trajectories, would be beneficial for the development of personalized treatments and patient care, improving outcomes and quality of life ^6,7,12^. The healthcare digital revolution can make this ambitious goal feasible, thanks to the increasing availability of detailed clinical and biological data in large population cohorts, that can be used for stratifying patients and develop predictive models of mental health outcomes. The UK Biobank (UKB) dataset is an example of this type of data, including information on a large cohort of about half million participants ^13^. At the same time, new analysis techniques driven by artificial intelligence (AI) and machine learning (ML) have emerged for handling high-dimensional clinical and biological data, to identify relevant biomarkers of complex disorders such as MDD, where contributions from both genetics and environmental factors interact to determine the disease and its outcomes ^14^.

In this framework, several unsupervised ML methods were employed to stratify MDD patients based on clinical ^15^, biological ^16^, and imaging data ^17,18^. However, the classical clustering algorithms, e.g., k-means clustering, present some limitations. Indeed, these data-driven ML algorithms would fail to stratify patients and thus extracting relevant subgroups if the structure of the data is large, complex and multidimensional ^17,19^.

In this context, new techniques as the data-analytics tool of Topological Data Analysis (TDA) represent valid options. TDA is a multidimensional data analysis tool, based on data topology geometry, which has the ability to reduce such high-dimensional data to simple and compact geometric structure (e.g., graphs), named topological skeleton, representing simple topological summary of the data from which can be directly captured fundamental characteristics of a dataset ^20,21^.

TDA is data driven and has the advantage of being able to capture the topological and geometrical structure of complex and large datasets, providing stability with respect to perturbations and noise in the input multidimensional dataset ^22^. TDA can successfully exploit higher order (more than pairwise) interactions among phenotypes, which are expected in genetic, environmental, clinical and brain imaging data, and provides outputs for rapid and intuitive exploration of the dataset structure, allowing to investigate how features converge or diverge in the defined clusters ^22^. Recently, TDA has been successfully employed in different medicine fields^20,23^ to navigate multimodal and high-dimensional biological datasets, including functional brain connectivity ^24^ and biomolecular structure ^25^, e.g. for the data-driven investigation of neuropsychiatric disorders such as attention-deficit/hyperactivity disorder (ADHD)^19^ and delirium ^22^ applied relatively on functional connectome and electroencephalograph (EEG) signals.

Beyond its applications to previously mentioned fields, TDA has proven to be also an efficient tool for augmenting and enhancing classical ML and deep learning methods defined as “topological machine learning” ^26^.

Of note, the joint application of TDA and Spatial Analysis of Functional Enrichment (SAFE) score analysis allowed to perform a multidimensional comparative analysis, enabling a quantitative characterization of MDD clinical outcomes stratification patterns based on different multimodal feature sets^27,28^. Specifically, SAFE score analysis has been applied to quantitatively examine the local enrichment created on the graph built on candidate predictive features by several functional outcomes^27^. SAFE score was demonstrated to provide a quantitative measure (e.g., through permutation) of statistical association between the network organization and selected outcomes ^27^, in contrast to most other methods, which mainly focus on the qualitative identification of subgroups within the TDA-based network by the visual inspection of the color-coded network obtained by mapping each node’s outcome ^23,26,29^.

In this framework, the multidimensional tool of TDA could drive a more precise stratification of MDD, leading to the identification of key biomarkers related to disease’s trajectories, such as TRD. For the first time, our study aimed to apply the novel multidimensional tool of TDA to provide a data-driven stratification of a large cohort of MDD from the UKB into meaningful clinical subtypes, by leveraging the unique UKB dataset.

A robust TDA pipeline was implemented and applied to genetic, environmental, and brain magnetic resonance imaging (MRI) features in UKB participants with MDD. TDA graphs built on each feature type and on their combinations were compared in terms of their capability to predict different clinical characteristics, measured as their degree of topological resemblance to the outcomes of interest. Based on prior methodological and clinical knowledge, we hypothesized that our TDA pipeline could effectively: (1) unveil intricate associations within complex and high-dimensional data from multiple sources, (2) provide new insights on MDD as heterogeneous disorder, specifically by uncovering previously unobserved MDD subgroups that exhibit differences in clinical outcomes supported by differences in the underlying multivariate predictive data.

## 2. Results

### 2.1 Participants characterization

After identifying the final group of MDD participants (n=3052) (Methods), different subsets were extracted based on the availability of information on different health-related outcomes of interest. In detail, three groups of outcomes were considered, relative to depression severity (Group 1, G1) (n=1861), cardio-metabolic and medical conditions (Group 2, G2) (n=3044), and TRD (Group 3, G3) (n=537). The clinical and socio-demographic characteristics of the MDD sample are shown in Table 1, considering the whole sample and split by the availability of each group of outcomes. *Figure 1* summarizes the demographic and clinical outcomes characteristics of participants included in this study.

**Figure 1.**
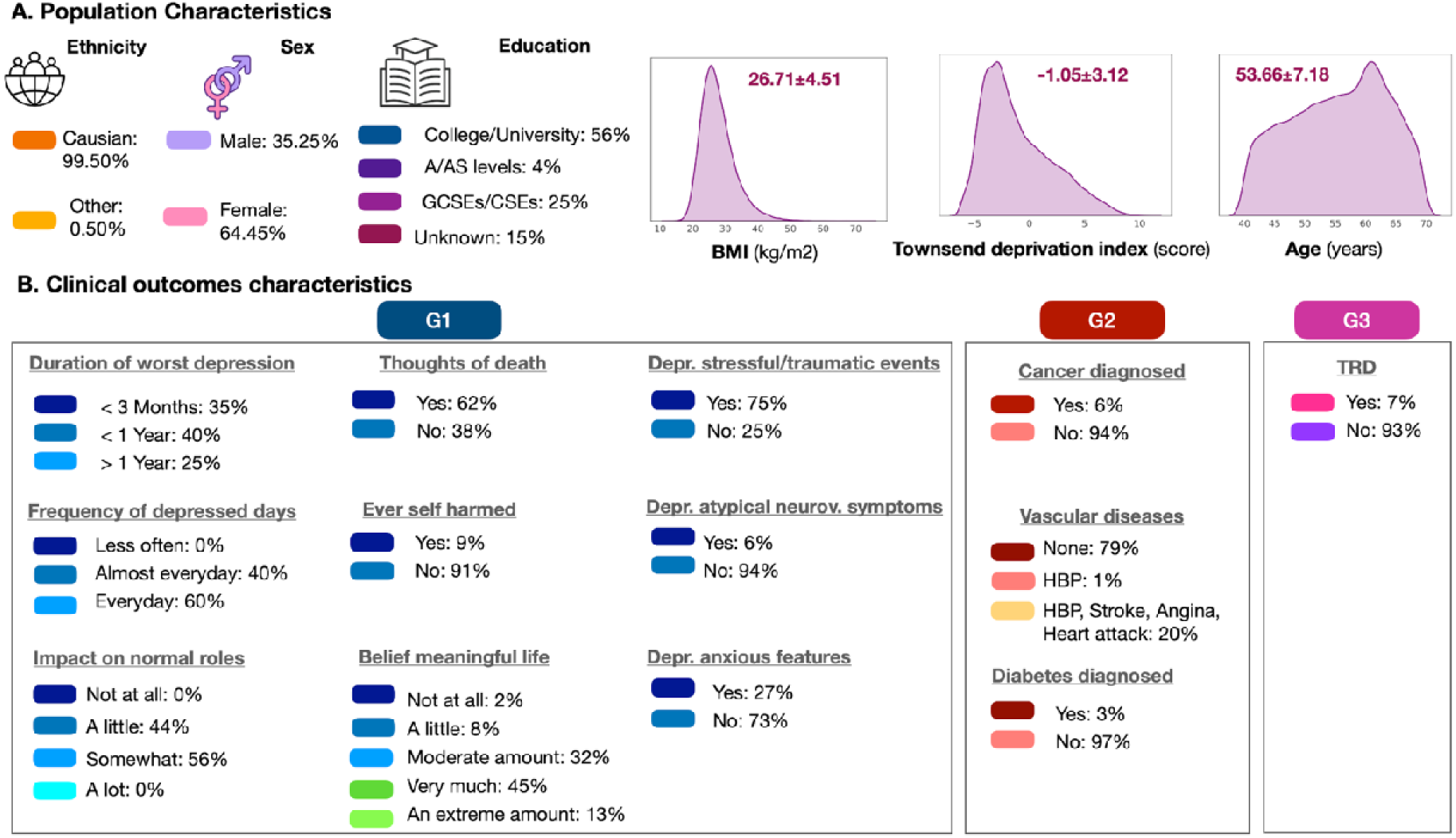
Socio-demographic (A) and clinical outcomes (B) characteristics for the participants included in the study. G1, clinical outcome group n. 1; G2, clinical outcome group n. 2; G3, clinical outcome group n. 3 (see also Table 5). Note that the number of individuals included in G1-3 varies as reported in Table 1, and that percentages reported in this figure were calculated considering the number of individuals in each outcome group and not the whole sample.

**Table 1.** Main clinical and socio-demographic characteristics of the sample. G1-G3 indicate each of the three considered groups of outcomes (see Table 2)

|  | Whole dataset | G1 | G2 | G3 |
| --- | --- | --- | --- | --- |
| **N subjects** | 3052 | 1861 | 3044 | 537 |
| **Sex<sup>a</sup>** |  |  |  |  |
| **Male** | 1076 (35.25%) | 611 (32.83%) | 1074 (35.28%) | 160 (29.79%) |
| **Female** | 1976 (64.45%) | 1250 (67.17%) | 1970 (64.71%) | 377 (70.20%) |
| **Age at baseline (years)<sup>b</sup>** | 53.66±7.18 | 53.27±7.05 | 53.66±7.19 | 53.63±7.27 |
| **BMI (kg/m<sup>2</sup>)<sup>b</sup>** | 26.71±4.51 | 26.90±4.68 | 26.71±4.51 | 26.76±4.54 |
| **WC (mm)<sup>b</sup>** | 86.91±12.64 | 87.14±12.97 | 86.93±12.75 | 86.54±13 |
| **Ethnicity<sup>a</sup>** |  |  |  |  |
| **Caucasian** | 3037 (99.50 %) | 1851 (99.46%) | 3029 (99.50%) | 536 (99.81%) |
| **Other ethnic background** | 15(0.50%) | 10 (0.54%) | 15 (0.50%) | 1 (0.19%) |

Abbreviations: BMI, Body mass index; WC, waist circumference. <sup>a</sup> Data expressed as count (percentage %). <sup>b</sup> Data expressed as mean ± standard deviation.

### 2.2 Pipeline Overview

The data analysis workflow is schematized in **Fig. 2**. From the group of MDD individuals included in our study (Methods), we extracted different subsets based on the availability of information of health-related outcomes of interest. Three groups of health-related outcomes were considered, relative to depression severity (n=1861), cardio-metabolic and medical conditions (n=3044), and TRD (n=537). We employed multimodal imaging and gene-environmental feature sets as input for the TDA to assess their differential capability to cluster MDD individuals into subgroups with homogeneous clinical outcomes. Specifically, multimodal MRI predictors included: T1-weighted structural MRI (sMRI), diffusion weighted MRI (dMRI), and resting-state (rs-fMRI) and task-based (t-fMRI) functional MRI data.

**Figure 2.**
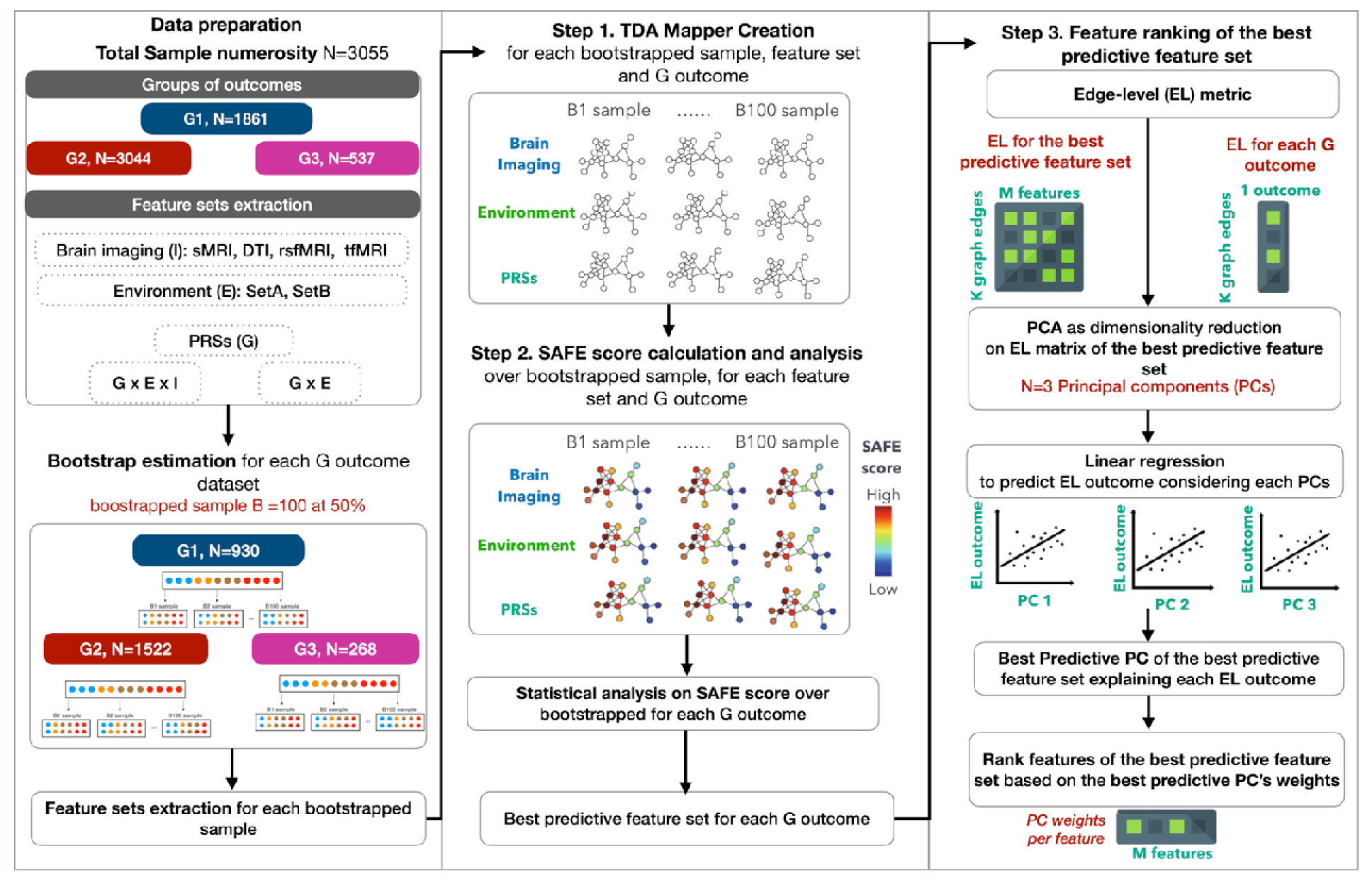
Data analysis framework. The steps include (from the left to the right panel): **Data preparation**, comprising division of the MDD individuals in three groups of health-related outcomes and feature sets extraction; **Bootstrapped samples estimation**, subdividing the feature set of each outcome’s group into 100 bootstrapped samples; **Feature sets extraction for the 100 bootstrapped samples** of each outcome’s group; **TDA Mapper application (step 1, Methods)**, creating 100 graphs for the 100 bootstrapped samples for the feature set of each outcome’s group; **SAFE score estimation and statistical analysis (step 2, Methods)** to extract the best predictive feature set for each outcome’s group; **Feature ranking (step 3, Methods)** comprising: the estimation of the EL metric, PCA application to the EL metric of the best predictive feature set, linear regression model application for the prediction of outcome’s EL metric to identify the best predictive PC, and ranking of the best predictive features based on the best predictive PC’s weights.

A bootstrap sampling without replacement (n=100) was applied to subdivide the feature set of each outcome’s group into 100 bootstrapped samples to build 100 graphs, and then compute 100 estimations of SAFE enriched scores for the selected clinical outcomes. As follow, the methodological steps (Methods) included: (step 1) TDA Mapper application, (step 2) SAFE score estimation and statistical analysis on its distribution over bootstrapped samples, (step 3) feature ranking which comprised: (a) estimation of the edge-level (EL) metric for the best predictive feature sets and for each outcome, (b) Principal Component Analysis (PCA) applied as dimensionality reduction step to the EL metric of the best predictive feature set, (c) Linear Regression applied for the prediction of EL metric of each outcome from the Principal Components (PCs) to identify the best predictive PC, (d) ranking of the features within the best predictive feature set based on the PC weights of the best predictive PC.

### 2.3 Graph-based outcome prediction

The bootstrapped TDA graphs built on the different feature sets were compared in terms of outcome predictive capability, defined as the SAFE score value found for each feature set representing the relevant association between features on which the network is constructed, and the health-related outcome analyzed (Methods).

After multiple comparison correction applied on Kruskal-Wallis (KW) tests, we showed differences among feature sets based on the SAFE score distributions over bootstrapped samples for all the outcomes (p<.005, surviving to Bonferroni correction). As shown in Table 3 and in Figures 3-4, depending on the outcomes, different feature sets resulted as best predictors according to the distribution of SAFE scores. Figures S1-S2 show the results of post-hoc pairwise comparisons performed for all the outcomes in the form of heatmaps of SAFE score differences among feature sets. For each outcome, the heatmap shows the significant differences in the mean ranks of SAFE score distribution between each pair of feature sets surviving multiple correction applied on KW test. The feature set (row) with the highest median value of SAFE score was considered the best predictor for the corresponding outcome (Figures S1-S2). For most outcomes in G1, feature sets including environmental variables were the best predictors (G-E-Imaging, G-E concatenation and environmental sets), while imaging-based feature sets were the best predictors for outcomes in G2 and G3 (Figures S1-S2).

**Figure 3.**
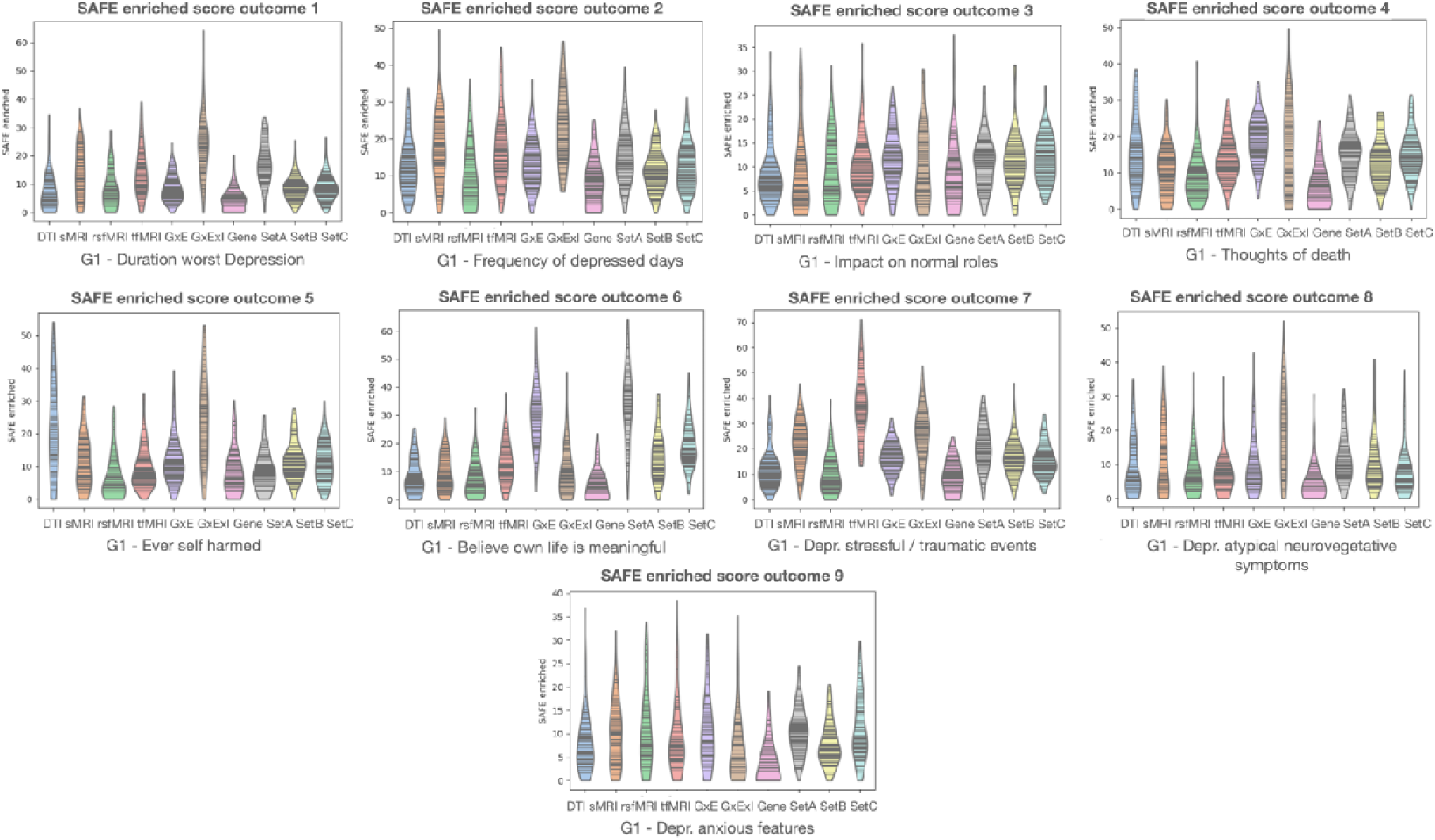
Violin plots of SAFE score distributions of all feature sets for G1 outcomes.

**Figure 4.**
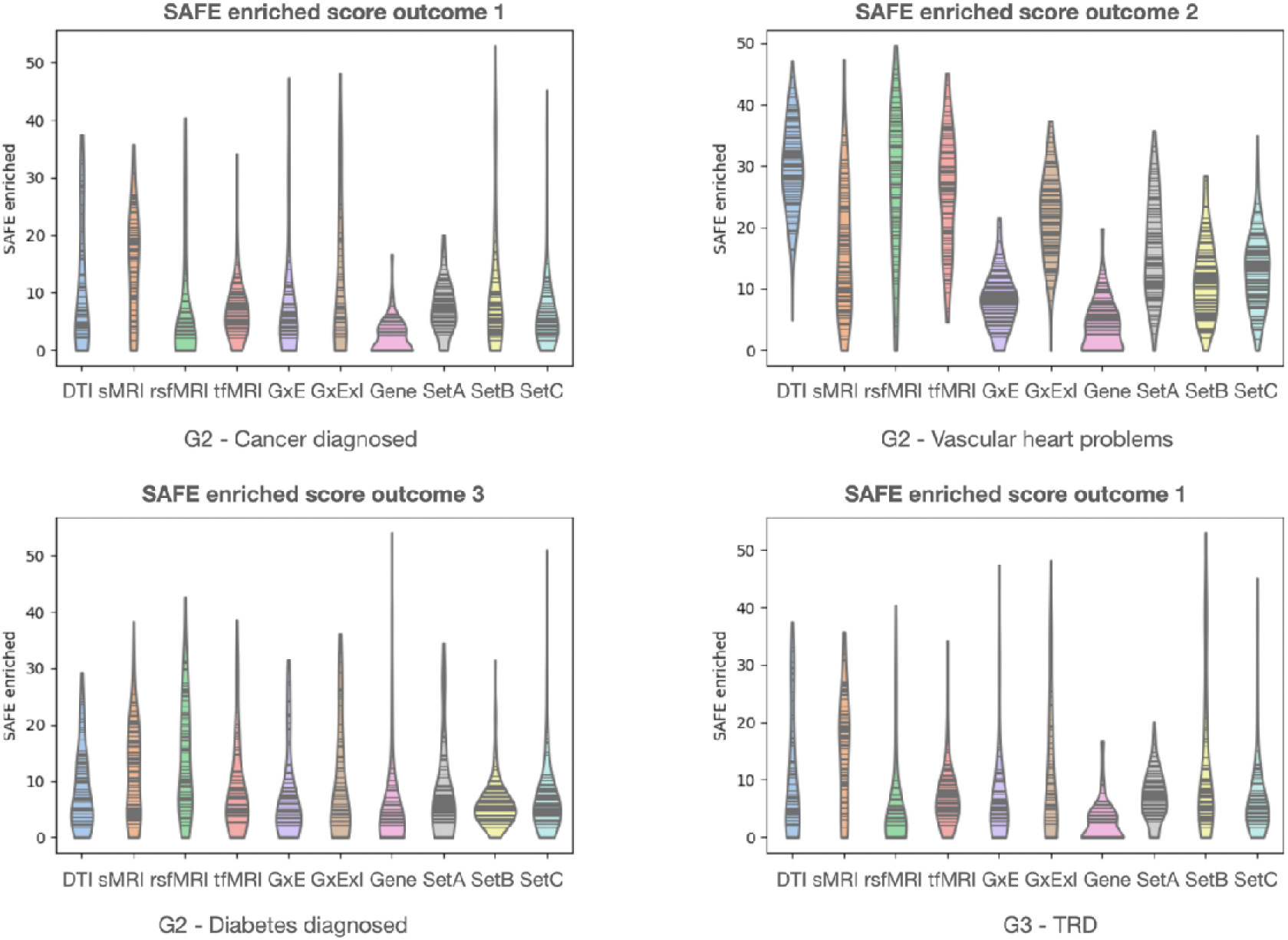
Violin plots of SAFE score distributions of all feature sets for G2 and G3 outcomes.

Post-hoc pairwise comparisons showed that for outcomes measuring the severity of MDD (worst depressive episodes, self-harming, and neurovegetative symptoms (O1, O2, O5, O8) the G-E-Imaging feature set emerged as the best predictor (i.e., the combination of imaging, genetic, and environmental variables).

Differently, for the outcomes measuring perceived life meaning and anxious symptoms (O6 and O9), the environmental SetA (features that were partly innate and partly affected by experience) was found as the best predictor. Environmental features (set C) reached the greatest SAFE score for the outcome referring to death thoughts (O4); further, the concatenation of these with genetic features (G-E feature set) resulted as the best predictors from SAFE score analysis for the outcome measuring the disease impact on normal roles (O3). Notably, stress-related depressive symptoms (O7) were best predicted by brain functional characteristics through the t-fMRI feature set.

For G2 outcomes, reflecting cardiometabolic and general health conditions, brain characteristics were more predictive than genetic and environmental features. Specifically, post-hoc pairwise comparison analysis showed that sMRI, DTI and rs-fMRI feature sets resulted the best predictors, associated with significantly higher SAFE scores than the sets composed of genetic and environmental features, for the outcomes cancer, vascular heart problems, and type 2 diabetes, respectively. As last, the t-fMRI feature set achieved the highest SAFE score for G3 TRD outcome, significantly higher with respect to the environmental and sMRI feature sets.

### 2.4 Feature ranking

We ranked features starting from the best predictive feature sets extracted from the SAFE score analysis, by firstly extracting the EL metric (Methods) for the best predictive feature sets and for each outcome, and as follows applying PCA to the EL metric of the best predictive feature set. Then, linear regression models were applied to extract the best predictive PC by considering as independent variable each PC with the aim to predict the EL metric of each outcome (Methods). As last, the ranking of the features within the best predictive feature set was performed by considering the PC weights of the best predictive one previously extracted (Methods).

The PCA results and the corresponding feature ranking for all the outcomes are summarized in Tables 2-3, including the best predictive PCs and related linear regression statistics for all the outcomes of interest. For all outcomes and the best PC, the top-five predicting features are reported in Table 2, sorted based on the magnitude of the PCA coefficient. Group-specific Bonferroni corrections were applied on the p-values of Table 2 (G1: p<.005, Bonferroni corrected for n=9, number of outcomes; G2: p<.01, Bonferroni corrected for n=3, number of outcomes; G3: p<.05, Bonferroni corrected for n=1, number of outcomes).

**Table 2.** , Best predictive PC, T-statistics and p-values from linear regressions considering as independent variable edge-related variation of the features of the best predictor to predict the edge-related variation of the outcome. Bold for p value surviving to group-specific Bonferroni correction (n=9 G1, n=3 G2, n=1 G3).

| G1 outcomes | Best predictor | PC1 |  | PC2 |  | PC3 |  |
| --- | --- | --- | --- | --- | --- | --- | --- |
|  |  | T-stat | p-value | T-stat | p-value | T-stat | p-value |
| O1 | GxExImaging | T=18.82 | **p<0.005** | T=-1.60 | p=0.1092 | T=4.07 | **p<0.005** |
| O2 | GxExImaging | T=31.37 | **p<0.005** | T=0.32 | p=0.75 | T=-3.02 | **p<0.005** |
| O3 | SetC Environment | T=16.66 | **p<0.005** | T=-1.00 | p=0.31 | T=6.18 | **p<0.005** |
| O4 | GxE | T=28.11 | **p<0.005** | T=1.48 | p=0.14 | T=1.52 | p=0.13 |
| O5 | GxExImaging | T=11.79 | **p<0.005** | T=1.29 | p=0.19 | T=-1.78 | p=0.07 |
| O6 | SetA Environment | T=14.93 | **p<0.005** | T=-2.18 | p<0.05 | T=-17.78 | **p<0.005** |
| O7 | tfMRI | T=17.00 | **p<0.005** | T=-7.99 | **p<0.005** | T=15.88 | **p<0.005** |
| O8 | GxExImaging | T=7.98 | **p<0.005** | T=-1.15 | p=0.25 | T=3.08 | **p<0.005** |
| O9 | SetA Environment | T=24.98 | **p<0.005** | T=-8.89 | **p<0.005** | T=-19.17 | **p<0.005** |
| **G2 outcomes** |  |  |  |  |  |  |  |
| O1 | sMRI | T=12.49 | **p<0.005** | T=2.03 | p<0.05 | T=-0.95 | p=0.34 |
| O2 | DTI | T=26.74 | **p<0.005** | T=-2.59 | **p<0.005** | T=1.90 | p=0.06 |
| O3 | rsfMRI | T=9.14 | **p<0.005** | T=-1.28 | p=0.20 | T=-0.71 | p=0.47 |
| **G3 outcome** |  |  |  |  |  |  |  |
| O1 | tfMRI | T=4.89 | **p<0.005** | T=-0.50 | P=0.62 | T=4.23 | **p<0.005** |

Abbreviations: G, group of clinical outcome; O, clinical outcome; PC, principal components; G, genetics; E, environment; sMRI, structural MRI; rsfMRI, resting-state fMRI; DTI, diffusion tensor imaging; tfMRI, task-based fMRI.

**Table 3.** Ranking of the five most important features extracted from the best predictive feature set for all outcomes. The most important feature in each predictive feature set is reported in bold.

| Predictive feature set for G1 outcomes | Feature ranking | PC coefficient |
| --- | --- | --- |
| **GxExImaging**<br>for O1, O2, O5, O8 | **Mean OD in Fornix** | 0.1477 |
|  | Mean ISOVF in left external capsule | 0.0908 |
|  | Mean RD in left superior fronto-occipital fasciculus | 0.0886 |
|  | Mean ISOVF in right external capsule | 0.0825 |
|  | Mean ISOVF in left superior fronto-occipital fasciculus | 0.0818 |
| **GxE**<br>for O4 | **Belittlement by partner or ex-partner as an adult** | 0.3142 |
|  | Felt hated by family member as a child | 0.2800 |
|  | Someone to take to doctor when needed as a child | 0.2442 |
|  | Physical violence by partner or ex-partner as an adult | 0.2385 |
|  | Childhood stressful events | 0.2354 |
| **SetA**<br>for O6, O9 | **Z adjusted T/S log (i.e., Telomeres length)** | 0.4784 |
|  | Summed MET minutes per week for all activity | 0.0626 |
|  | Ethnicity | 0.0070 |
|  | Able to confide | -0.0296 |
|  | Frequency of friend or family visits | -0.0817 |
| **tfMRI**<br>for O7 | **Median z-statistic (in group-defined mask) for shapes activation** | 0.3882 |
|  | 90th percentile of z-statistic (in group-defined mask) for shapes activation | 0.3849 |
|  | 90th percentile of z-statistic (in group-defined amygdala activation mask) for faces-shapes contrast | 0.3729 |
|  | Median z-statistic (in group-defined amygdala activation mask) for faces-shapes contrast | 0.3579 |
|  | Median z-statistic (in group-defined mask) for faces-shapes contrast | 0.3112 |
| **SetC**<br>For O3 | **Felt hated by family member as a child** | 0.4419 |
|  | Be littlement by partner or ex-partner | 0.4036 |
|  | Childhood stressful events | 0.3799 |
|  | Someone to take to the doctor when needed as a child | 0.3169 |
|  | Physically abused by family as a child | 0.2898 |
| **Predictive feature set for G2 outcomes** | **Feature ranking** | **PC coefficient** |
| sMRI<br>for O1 | **Volume of parsopercularis (right hemisphere)** | 0.1437 |
|  | Area of parsoeprcularis (right hemisphere) | 0.1348 |
|  | Area of supramarginal (right hemisphere) | 0.1180 |
|  | Mean thickness of precentral (right hemisphere) | 0.1073 |
|  | Volume of middle temporal (left hemisphere) | 0.1059 |
| DTI<br>for O2 | **Mean OD in fornix** | 0.3516 |
|  | Mean RD in left superior fronto-occipital fasciculus | 0.1874 |
|  | Mean RD in fornix | 0.1405 |
|  | Mean RD in right superior fronto-occipital fasciculus | 0.1347 |
|  | Mean ICVF in left superior fronto-occipital fasciculus | 0.1166 |
| rsfMRI<br>for O3 | **Strength of negative weights in IC 54 (Sub&Cereb)** | 0.1496 |
|  | Strength of negative weights in IC 30 (SMN) | 0.1384 |
|  | Strength of positive weights in IC 11(DMNp) | 0.1347 |
|  | Strength of positive weights in IC 2 (VAN) | 0.1328 |
|  | Strength of positive weights in IC 53 (Sub&Cereb) | 0.1306 |
| **Predictive feature set for G3 outcome** | **Feature ranking** | **PC coefficient** |
| tfMRI<br>for TRD | **Median z-statistic (in group-defined mask) for faces-shapes contrast** | 0.4657 |
|  | 90th percentile of z-statistic (in group-defined amygdala activation mask) for faces-shapes contrast | 0.4362 |
|  | Median z-statistic (in group-defined amygdala activation mask) for faces-shapes contrast | 0.3744 |
|  | 90th percentile of z-statistic (in group-defined mask) for faces-shapes contrast | 0.3285 |
|  | 90th percentile of z-statistic (in group-defined mask) for shapes activation | 0.3149 |

Abbreviations: G, group of clinical outcome; O, clinical outcome; PC, principal components; G, genetics; E, environment; sMRI, structural MRI; rsfMRI, resting-state fMRI; DTI, diffusion tensor imaging; tfMRI, task-based fMRI; OD, orientation dispersion index; ISOVF, Isotropic Compartment Volume Fraction; RD, radial diffusivity; MET, metabolic equivalent task; ICVF, Intracellular Volume Fraction; SMN, somatomotor network; DMNp, default mode network posterior; VAN, ventral attention network; Sub&Cereb, subcortical and cerebellar network.

PC1 resulted as the best predictive one for all the outcomes, except for the G1 outcome relative to perceived life meaning (O6), for which PC3 was associated with the highest T statistics in the linear regression.

G-E-Imaging feature set resulted the best predictor for the majority of G1 outcomes (40% of G1 outcomes), especially the ones measuring worst depressive episodes, self-harming, and neurovegetative symptoms (O1, O2, O5, O8). Within the G-E-Imaging feature sets, the DTI feature of mean ODI in the fornix resulted the most important.

As follows, environmental sets (i.e., SetA and SetC) resulted still relevant for the 30% of G1 outcomes, especially for the ones related to perceived life meaning, anxious symptoms, and outcome concerning disease impact on normal roles (O3, O6, O9). Precisely, the feature telomeres length from environmental SetA and the variable “felt hated by family members as a child” from environmental SetC resulted the most relevant.

Further, brain imaging predictors were reported as the most important feature sets for medical comorbidities outcomes and TRD. Specifically, the structure brain characteristic of volume of pars opercularis (right emisphere) was relevant for cancer (O1 for G2), whereas rs-fMRI feature of strength of negative weights in subcortical and cerebellar network (Independent Component (IC) 54) and DTI feature of mean OD in fornix were relatively relevant for vascular heart problems and diabetic outcome (O3 for G2). Among functional brain characteristics relevant for TRD (G3), the t-fMRI median z-statistic for faces-shapes contrast resulted the most important features.

## 3. Discussion

In the present work, we reported the novel application of TDA to assess the differential performance of multimodal datasets including genetic, environmental, and brain characteristics to stratify individuals with MDD based on different health outcomes, embracing disease severity, medical comorbidities, and TRD. For the first time, TDA was employed to perform a multivariate stratification of MDD based on diverse sets of predictors on the large UKB cohort, enabling a comprehensive, robust, and data-driven exploration of this heterogeneous disease.

The data-driven TDA approach revealed a complex picture of predictors, unraveling the multitude of domains that contribute to stratify MDD based on the selected clinical outcomes. Consistent with previous knowledge ^14,30–32^, we found a key role for the environment, alone or integrated with biological factors, in determining the severity of depressive symptoms. Conversely, brain characteristics emerged as the most relevant predictors for medical comorbidities and TRD.

Overall, these findings support the hypothesis that the consideration of genetic, environmental, and brain characteristics is essential to characterize the heterogeneity of MDD phenotypes. TDA has emerged as a promising unsupervised tool for identifying candidate biological and environmental markers of disease outcomes based on different multimodal feature sets, that could be used to predict disease-related trajectories. If replicated on independent cohorts, our results will set the ground for a meaningful dimensional stratification of MDD, for clarifying the corresponding biological and environmental underpinnings, and potentially for developing clinical applications.

### 3.1 Multivariate data analysis of TDA and application of SAFE score

In this study, we applied TDA to different multidimensional feature spaces, for the first time in a large cohort of MDD individuals from the UKB. TDA applies mathematical concepts from geometric topology to characterize the shape of a multidimensional dataset into a simple and compact geometric structure, named topological skeleton, representing a simple topological summary of the data. This approach allows the investigation of multidimensional data once in a step, and further the characterization of the dataset in terms of geometrical and topological information extracted from the topological skeleton^26^. Examples of TDA in precision medicine context are beginning to appear in the literature, including the definition of clinically meaningful phenotypes for diverse medical conditions^19,20,23^. However, TDA was never applied before to multimodal (genetic, environmental, neuroimaging) and high-dimensional datasets in an MDD population. Therefore, the availability of quantitative functional maps created by the SAFE score allows an increase in the interpretability of the association between features on which the network is constructed and the target outcome analyzed, especially in the case of high-dimensional complex networks for which the network’s visual inspection could be computationally challenging. Given that clinical applications are high-stakes, we require understandability from the prediction tools or either they will grow in distrust^33^. This is especially true in the analysis of the neurobiological underpinnings of psychiatric disorders, where innovative statistical tools should assist clinicians without introducing further complexity, by proving to be trustworthy, therefore not only valid and reliable, but also easily understandable. Therefore, the TDA technique, coupled with the extraction of SAFE score functional maps, provides the simultaneous possibility of multifactorial data aggregation, capturing the variability of multiple feature sets, and an improvement in interpreting the associations between TDA networks and target outcome variables.

### 3.2 Environment as a key determinant of depressive symptoms

We observed a key influence of the environment on depressive symptoms, as it represented the most predictive feature sets for most G1 outcomes, either alone or in combination with genetic and brain features.

Specifically, intertwining complex effects of biological and environmental features were observed for the outcomes duration of worst depressive episode and self-harm behaviors. It is established that the pathogenesis of MDD can be traced back to the synergic interaction between biological aspects (e.g., genes and neurobiology) and environmental risk factors^30^, although the relative weight of the single factors is unclear. Apart from the etiopathogenesis, recent studies suggested that outcome measures, such as global functioning, could be predicted using a combination of baseline clinical, brain morphological, and environmental information^14^. A direct link has also been established between various environmental factors and brain structural and functional changes, highly correlated with severe depressive symptoms and poor prognosis^31,32^.

Brain characteristics might therefore act as mediators of the environment. Consistently with this hypothesis, within the G-E-Imaging feature set of predictors, the microstructural integrity of the fornix was one of the most relevant predicting features for G1 (severity-related) outcomes. This is in line with previous studies suggesting that the fornix and stria terminalis are involved in the pathophysiology of mood and psychotic disorders^34,35^. White matter (WM) abnormalities in the fornix have been described in patients with early-onset MDD ^36^, as well as during late-life depression^37^. Further, alterations in the fornix in patients with MDD might impair connections between brain regions such as the hippocampus and prefrontal cortex, which are important for depression and psychological functioning ^38^.

Depression-related outcomes reflecting anxiety, life meaning, and suicidal ideation, were best explained by environmental factors, with a key role of telomere length, reflecting both innate and environmental characteristics. Conversely, childhood stressful events, especially the feeling of being hated by family members as a child, were associated with the impact of the disease on normal roles. Previous studies have investigated the impact of the environment on the severity of depression, identifying early life adversities, stressful life events, socioeconomic status, and exposure to traumatic events as determinants of the progression and intensity of depressive symptoms ^31,32,39,40^.

Finally, regarding telomere length, this feature has been previously associated with psychiatric disorder and especially MDD^41,42^. Telomere length can be considered a cellular clock, affecting how quickly cells reach senescence^43^. A compromised telomere biology has been associated with different medical conditions and recent studies suggested telomere shortening as a potential mechanism by which MDD may increase the risk of morbidity and mortality^41^. The causal nature of this association is not known; recent literature suggested an interaction between inflammation and telomeres, with the possible mediation of gut microbiome^42^. Telomere shortening is known to result from repeated mitotic divisions and exposure to a variety of cellular stress mechanisms^44^. Thus, activation of telomerase activity during stress may represent one of the compensatory mechanisms to withstand stress-related disorders. In this sense, it has been speculated that MDD is associated with increased cellular stress and replication, resulting therefore in accelerated telomere shortening^45^. However, more studies are needed to understand the prospective importance of telomere length in MDD. Particularly, mixed results are reported regarding the association between telomere shortening and severity of MDD, with some studies reporting a link between telomeres and severity measures ^46^, while others reporting negative results^47^.

Taken together, these findings highlight the importance of considering the role of the environment in the assessment of MDD, as it can shape disease severity and potentially progression. Our results confirm that the integration of environmental factors within a predictive stratification framework might improve outcome prediction.

### 3.3 Brain imaging predictors for medical comorbidities of MDD

Altered connectivity in selective brain regions, abnormal structural brain measures, in combination with lifestyle factors and chronic stress events, were previously associated with clinical outcomes of medical comorbidities conditions^48^.

Our results suggest that comorbid medical conditions in MDD could be better predicted by brain imaging features than by genetic and environmental ones. Specifically, the best predictors for cancer, type 2 diabetes, and vascular diseases were sMRI, DTI, and fMRI feature sets, respectively. Previous evidence of a role of brain features in shaping cardiometabolic risk factors is limited. Although different studies analyzed the link between depressive disorders and cardiometabolic diseases^49^, the brain underpinnings of this link are less known. The autonomic regulation and specifically the dynamics related to central-autonomic network (CAN) is one of the most studied mechanism that mediates the link between depression and cardiovascular health^50,51^.

However, other studies provided insights on the association between alterations in neural substrates and cardiometabolic conditions in the general population (e.g., diabetes, cardiovascular diseases/events, cancer, autoimmune disease), as well as between altered brain patterns and the risk of these conditions. Independent associations between cardio- and cerebrovascular risk factors and brain imaging changes were found to anticipate the disease manifestation^52^. On the other hand, only few studies suggest an association of neuropsychiatric disorders, including MDD, with medical conditions and concomitant alterations in brain features^48,53,54^. Indeed, the specific role of brain patterns, including predictors, moderators, or mediators for such clinical factors, remains unexplored. Our results suggest different brain imaging predictors for the different comorbidities examined, with possible roles of frontal morphology in cancer, fornix microstructure in cardio-vascular problems, and functional connectivity in the subcortical and cerebellar network in type 2 diabetes.

In conclusion, our understanding of the role of brain imaging features in general medical comorbidities of MDD remains largely incomplete, and mostly limited to the study of CAN dysfunctions^55,56^. Further studies are needed to clarify the pathways connecting depressive disorders, general health diseases, and brain characteristics.

### 3.4 fMRI features as predictors of TRD

Previous insights highlighted the complex interplay between environmental factors and brain features in defining other challenging clinical outcomes, such as TRD^57^. Indeed, specific MRI features related to brain structure and function have been reported as possible markers of TRD, demonstrating structural abnormalities and disrupted connectivity within critical brain regions of frontolimbic areas, including prefrontal, anterior cingulate cortex, hippocampus, amygdala and insula observed in patients with MDD and poor treatment outcomes^58,59^. Our data showed that brain responses to emotional-cognitive tasks were the best predictors for TRD in the UKB. Specifically, the most important feature associated with TRD was the brain activation elicited by faces with negative emotions. Recent literature^60^ suggested that brain function captured by fMRI might differentiate TRD when compared both to healthy controls and to MDD patients who respond to treatment, especially in emotional and reward brain areas. In particular, an alteration in amygdala response to emotional processing has been reported^61^. Consistently, a decreased ventromedial and ventrolateral prefrontal-amygdala connectivity during face processing seems to be reversed by the amelioration of symptoms following treatment with psilocybin or administration of selective serotonin reuptake inhibitors^62^. Although the brain imaging features used in our study were not analyzed in relation to TRD by previous studies, our findings remark that fMRI aspects might be important to discriminate TRD and predict treatment response.

However, the understanding of the risk factors of TRD remains limited for several reasons. First, the difficulty to enroll large samples characterized for TRD in fMRI studies, which was partly overcome in our study compared to previous ones^60^. Second, TRD samples may have an increased heterogeneity vs overall MDD, due to the long-term treatment of these patients with multiple medications and the different pathways that might lead to TRD. In this context, TDA provides the advantage of simultaneously handling all the variables in a common multifactorial space that reflects the structure of underlying dataset.

### 3.5 Limitations

This study presents potential limitations that need to be discussed. First, TDA application is not straightforward and requires the tuning of multiple parameters. In our study, the resolution and gain TDA parameters have been defined by varying each metric within a range and, through visual inspection, by choosing the ones that ensure that the majority of subjects is included in a connected node, and all the nodes are connected. In future applications, a methodological pipeline for defining TDA parameters is desirable. The graph that is obtained from the high-dimensional raw dataset is highly sensitive to the definition of gain and resolution, as the size and the overlap between bins in the filtered space are responsible for the definition of a coarse-grained network. Similarly, the choice of the filter function, number of filter’s projections, and clustering method might have influenced the results.

Specifically, a comprehensive exploration of covering parameters’ combinations, as well as filtering and clustering methods, might be performed by using as quality metrics the stability and connectivity measures of the network. Nevertheless, although a full exploration of Mapper’s parameters is necessary, the hyperparameter tuning could still depend on arbitrarily choices made by the user or selections based on the specific application (i.e., range of parameters to be explored), thus indicating either way a still arguable solution, not completely free from user’s choice. However, our approach currently represents the most common methodology for TDA parameters setup^20^.

Secondly, the selection of the predictive and outcome features inevitably entailed choices that might be questioned, as well as the definition of outcomes such as TRD, which was derived from the number of antidepressant switches within certain time frames according to primary care records, as previously described^63^. Among the candidate predictors, a delicate choice regarded the inclusion of pharmacological therapy as a candidate confounding predictive factor in all feature sets, which was motivated by the hypothesized interaction of therapy with all the other sets. Further, the reason for taking medications (i.e., disease diagnosis), the duration, the dosage, the response and/or adverse effects related to antidepressant medications were not considered, thus impeding further analysis with actual treatment response and dosage of medications.

As follow, the methodological choice related to the number of bootstrapped iterations (n=100) used for evaluating the significant association patterns among graph features-outcomes could have represented a limit for the analysis. Thus, further studies might employ higher number of bootstrapped iterations, trying to balance the identification of a stable SAFE score estimation and computational resources to create a complex graph on a large dataset. Moreover, an external set for validation is required and further studies are needed to replicate our results within an external validation set.

Moreover, although SAFE score application within Mapper context introduced by *Liao and colleagues* represents a valuable first metric to quantify statistical associations between network organization and selected outcome variables, future studies introducing new metrics to investigate Mapper graph’s feature-outcome association are needed. Indeed, besides defining the graph’s feature-outcome association at the level of the node, as is done by SAFE analysis, a full characterization of the graph’s topological properties might be useful for further exploitation, with the aim to develop new integrative framework based on TDA for several disorders’ stratification.

In addition, the classification of environmental factors in two main classes used in this study was meant to simplify the input data and increase interpretability, however, other classifications could have been applied. Among brain features, our study considered as functional features of resting-state the strength of positive and negative connections of the brain networks identified through ICA. Alternative topological measures of the functional brain networks, such as degree, centrality, clustering coefficients, and modularity, could have been considered.

To conclude, the MDD individuals considered in this study may not be representative of the general MDD population. Indeed, UKB is known to be enriched in females, elderly, wealthier and more educated individuals vs the general UK population^64^.

## Conclusions

Our study has exploited TDA as a powerful approach to investigate the biological and environmental markers of several outcome domains in a large MDD population, spanning from disease severity to medical comorbidities and TRD.

We highlighted key roles of the environment in MDD severity, brain features in medical comorbidities such as cardio-metabolic diseases, and functional brain features in TRD. Our findings suggested that multivariate data analysis based on data-driven TDA enables the handling of high-dimensional datasets and the extraction of hidden relationships among multiple types of features. Of note, the application of SAFE score analysis within a TDA pipeline enables a quantitative understanding of how TDA networks are functionally organized with respect to a specific target outcome.

Despite the need to test our findings on independent samples, this study provides avenues for the robust definition of biologically- and environmentally-determined dimensions affecting relevant health outcomes in MDD.

## Supporting information

https://osf.io/2se7r/?view_only=9cc533ad4c05449dabfd8613ad9dfdf7

## 4. Materials and Methods

### 4.1 Participants

The UKB is a population-based cohort from the United Kingdom, including ∼500,000 individuals. UKB has collected longitudinal environmental, lifestyle, activity, genetic, multimodal neuroimaging data, and other biomarkers, as well as health-related information^13^. We included in this study participants with a diagnosis of MDD according to any measure/assessments available. In detail, we considered: 1) diagnosis of a depressive disorder according to primary care records, considering diagnostic codes previously described (at least one diagnostic code for depression, n=37,394)^63^; 2) MDD defined by the Composite International Diagnostic Interview Short Form (CIDI-SF) (n=34,870), which was part of the Mental Health Questionnaire (MHQ) ^65^; 3) hospital diagnoses (ICD-10 codes F32-F33) (n= 17,271); 4) Smith et al. definition (n=30,876)^66^. Participants with a diagnosis of a bipolar, psychotic or substance use disorder according to primary care records, hospital ICD-10 codes, and/or MHQ data were excluded, as described also in ^63^. In total, we included 95,741 individuals with lifetime MDD according to one or more of the described criteria. Among these individuals, 3,052 participants were selected based on the availability of information on all outcome predictors, i.e., socio-demographic, genetic, environmental, and multimodal imaging measures. From this group of participants, different subsets were extracted based on the availability of information on different health-related outcomes of interest. Specifically, the three groups of health-related outcomes were considered, relative to depression severity (n=1861), cardio-metabolic and medical conditions (n=3044), and TRD (n=537).

### 4.2 Measures

#### Brain imaging predictors

Multimodal MRI information available from the UKB was extracted, considered as candidate predictor of MDD characteristics, and included as input for the TDA. Pre-processing steps and quality control analyses applied to extract the following brain imaging characteristics were previously described in literature^67^.

The dataset included T1-weighted sMRI, dMRI, and rs-fMRI and t-fMRI data relative to an implicit emotion processing task^68^. Specifically, sMRI features (n=200) included 14 subcortical volumes (SubV), 62 cortical gray matter volumes (GMV), 62 cortical thickness (CT), and 62 surface areas (SA) extracted from the Desikan-Killiany atlas regions of interest (ROIs). The DTI features (n=384) included fractional anisotropy (FA), mean diffusivity (MD), orientation dispersion index (OD), Intracellular Volume Fraction (ICVF), Isotropic Compartment Volume Fraction (ISOVF), axial diffusivity (AD), radial diffusivity (RD), and L1 direction extracted from the 48 tract ROIs defined using the Johns Hopkins University tract atlas.

The rs-fMRI features (n=110) consisted of the strengths of positive and negative weights extracted from the 55 x 55 adjacency functional connectivity (FC) matrix among pairs of non-artificial group-level spatial ICs. Each of the 55 ICs obtained from group ICA were grouped in nine resting-state functional networks (RSNs) reported in Table S1, in accordance with previous studies^69^. As last, n=8 summary measures of t-fMRI activations were extracted, including the 90th percentile of BOLD effect (4 measures) and of the z-statistic (4 measures) for faces-shapes contrast, faces activation, and shapes activation in group-defined mask, and for faces-shapes contrast in group-defined amygdala activation mask. Details on the MRI data pre-processing and feature extraction steps are reported in the **Supplementary Materials**.

#### Genetic-environmental predictors

The following genetic-environmental information was included as input in the TDA.

***Environment***. Environmental features, listed in Table 4, were either divided into two feature sets (SetA and SetB) or considered together (SetC). A detailed description of each environmental variable within SetA and SetB is reported in Table S3 and Table S4. SetB covered solely the environmental factors “experienced” by participants during their lifetime, including substance use (i.e., alcohol, smoking, and cannabis), lifestyle related to dietary changes, and traumatic or stressful events. SetA included characteristics partly innate and partly resulting from lifetime events, such as personality traits, social support, telomere length (corrected for the confounding factor of total white cell counts^70^, chronotype, and physical activity. SetC was defined as the concatenation of SetA and SetB. A detailed description of the variables included in each set is reported in Tables S3 and S4.

**Table 4.**
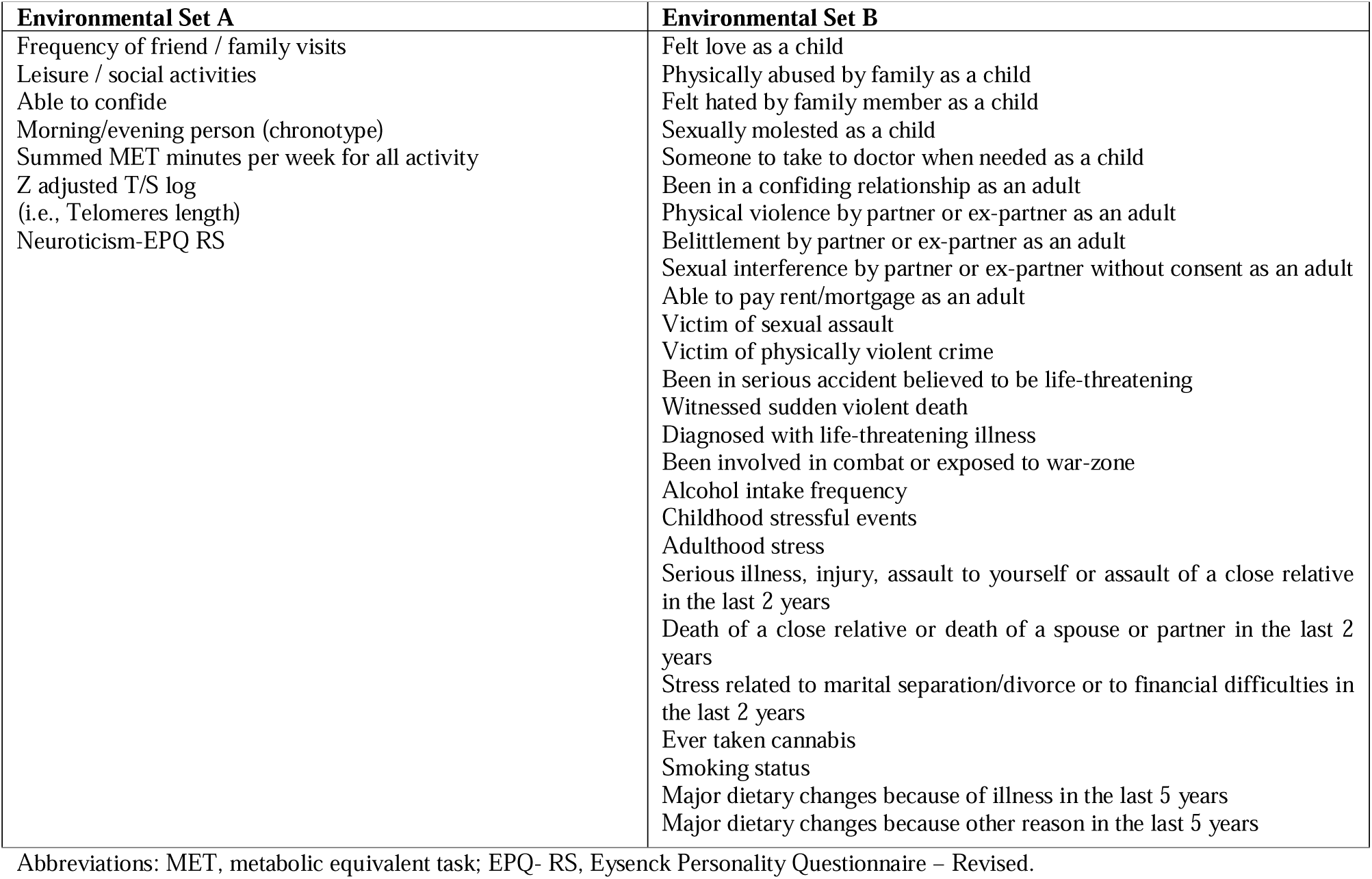
Environmental characteristics included in SetA and SetB.

***Genetics***. Genetic features consisted of polygenic risk scores (PRSs) estimated using PRS-CS-auto^71^ and calculated using the score function in PLINK 2.0^72^ in participants of European ancestry (defined by 4-means clustering on the first two genetic principal components^63^. We considered the PRSs of both psychiatric and non-psychiatric traits (see Table S2 for a description of the GWAS summary statistics used). In detail, we estimated the PRSs of major psychiatric disorders (bipolar disorder, schizophrenia, MDD, anorexia nervosa, autism, attention deficit hyperactivity disorder, alcohol dependence), related PRSs (neuroticism, smoking, alcohol consumption, years of education), and PRSs of immune-cardiometabolic traits (C-reactive protein, glycated hemoglobin, triglycerides, LDL and HDL cholesterol, coronary artery disease, body mass index, type 2 diabetes mellitus). The PRS of immune-cardiometabolic traits were considered given their clinical and pathogenetic correlation with MDD^73^, also in relation to the inclusion of cardio-metabolic comorbidities among the outcomes of interest. Each PRS was adjusted for ancestry-relevant population principal components, genotyping batch and centre of recruitment before inclusion in the TDA.

#### Confounding variables

Age, sex, ethnicity, and use of antidepressant medication(s) were considered as confounding input features for the TDA. The variable of ethnicity was converted into a dichotomous variable (1, Caucasian; 0 Other ethnicity). Socio-demographical variables, including Age, sex, ethnicity, are described in Table S6.

The considered antidepressant medications are described in Table S7. A dichotomous treatment indicator was extracted (i.e., 1 for participants taking any antidepressant medication, 0 for the other participants).

#### Health-related outcomes

Information on physical and mental health was used to characterize MDD subgroups in terms of clinically relevant outcomes. The selected features were divided into three outcomes groups, as reported in Table 5. A detailed description of each outcome variable is reported in Table S5.

**Table 5.**
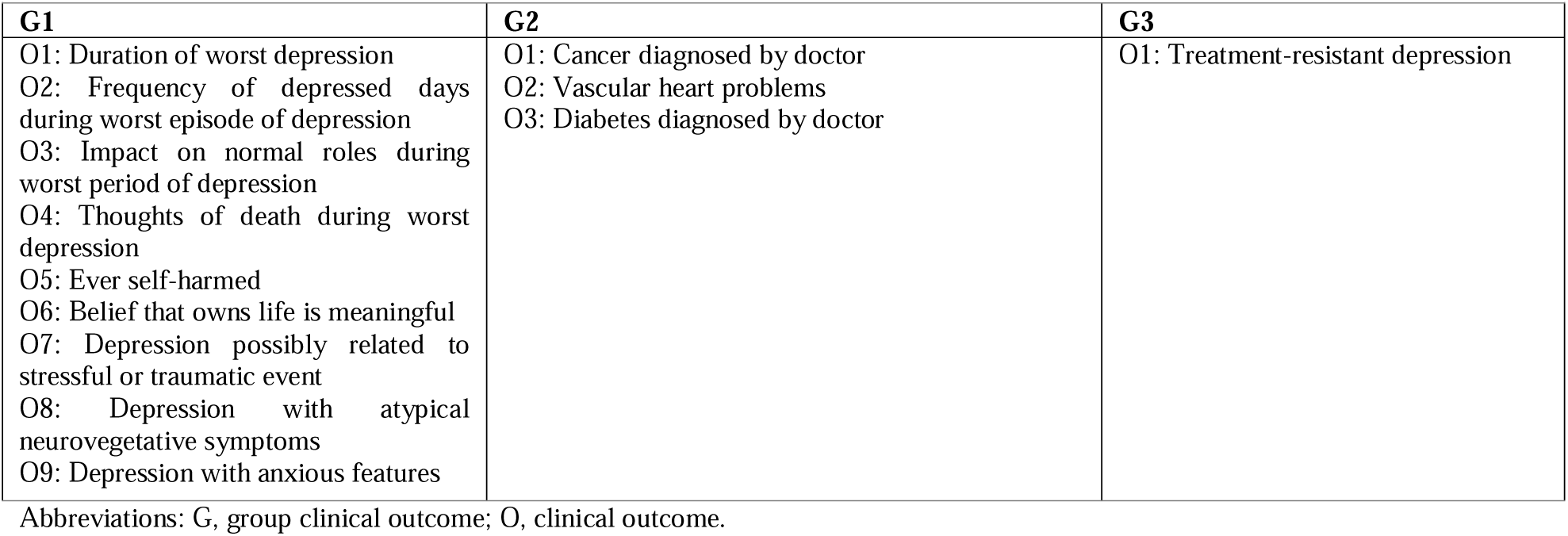
Description of clinically relevant outcomes considered in each group.

| **G1** | **G2** | **G3** |
| --- | --- | --- |
| O1: Duration of worst depression<br>O2: Frequency of depressed days during worst episode of depression<br>O3: Impact on normal roles during worst period of depression<br>O4: Thoughts of death during worst depression<br>O5: Ever self-harmed<br>O6: Belief that owns life is meaningful<br>O7: Depression possibly related to stressful or traumatic event<br>O8: Depression with atypical neurovegetative symptoms<br>O9: Depression with anxious features | O1: Cancer diagnosed by doctor<br>O2: Vascular heart problems<br>O3: Diabetes diagnosed by doctor | O1: Treatment-resistant depression |

Abbreviations: G, group clinical outcome; O, clinical outcome.

The clinical outcomes were divided in: i) variables related to severity of depression (i.e., type and duration of depression, self-harm behaviors, as well as depression with anxious and neurovegetative symptoms defined as in ^74,75^)(**G1)**; ii) variables related to cardio-metabolic and general health conditions (i.e., cancer, type 2 diabetes, cardiovascular diseases) (**G2)**; iii) TRD, defined as having at least two switches between different antidepressant drugs (independently on the class) (**G3**), as detailed in a previous work^63^.

### 4.3 Data Analysis

Multimodal imaging and gene-environment feature sets were employed as inputs for the TDA to assess their differential capability to cluster MDD individuals into subgroups with homogeneous clinical outcomes. We used TDA Mapper, provided by the *tmap* library^28^. Most previous TDA applications assessed graph-outcome associations based on a qualitative basis^23,26,29^. On the other hand, the SAFE score allows a quantitative estimation, previously described in the applications of Liao and collegues^28^ and Baryshnikova and collegues^27^. The SAFE score maps the values of a target variable of interest onto the Mapper graph and extracts significant association patterns, called “subgraphs enrichment”, for the specific target variable. This score represents the only quantitative metric used in the literature for assessing the outcome predictive capability of a TDA graph^27,28^.

#### TDA Mapper application: Step 1

Different graphs were built by separately employing the different feature sets, as follows: (i) sMRI features, (ii) DTI features, (iii) t-fMRI features; (iv) rs-fMRI features; (v) genetic features (G); environmental features including (vi) SetA, (vii) SetB, (viii) SetC; (ix) genetic-environmental (SetC) features (G-E); (x) genetic-environmental-imaging (G-E-Imaging) features. Confounding features were added as inputs to all feature sets, to estimate their predictive capability at the net of these confounding variables. For each of the feature sets, three graphs were built for the three outcomes’ groups (G1, G2, G3) identified before.

In our application, the subjects’ space S was defined as the selected feature set. The Euclidean distance was chosen as the metric of distance (i.e., similarity) between data points in the subjects’ space S. As a data preparation step, the data matrix of each feature set was standardized according to the robust data scaling. The original patterns were projected along two dimensions using Uniform Manifold Approximation and Projection for dimensionality reduction (UMAP) with default parameters. The mean and variance of the projected data matrix was computed to assess the variance-normalized Euclidean distance metric between projected data points. A bootstrap sampling without replacement (n=100) was applied, to build 100 graphs, and then compute 100 estimations of SAFE enriched scores for the selected clinical outcomes. The bootstrap percentage was set to 50%, indicating that half of all the subjects were chosen from the original dataset to form a resampling subset (n=930 subjects for G1, n=1522 subjects for G2, n=268 subjects for G3). Specifically, we subdivided the feature set of each outcome’s group into 100 bootstrapped samples, choosing a dimension of 50% of the original dataset. The number of bootstrapped resampling was chosen balancing the extraction of a complex graph construction (i.e., large number of features and samples), SAFE score estimation and computational time of our application. Thus, we wanted the algorithm to run in a reasonable amount of time on large graphs (i.e., considering large number of features and samples), considering that the computational complexity of graph’s construction remains high for a large graph^76^. As following step, the filtered data were divided into overlapping intervals defining a cover, that was characterized by the parameters of Resolution (R, number of overlapping bins) and Gain (G, overlap between bins). In our case, cover parameters were selected by testing a wide range of R-G pairs on the G-E-Imaging feature set (one per outcome group); based on visual inspection, R=15 and G=4 pair was selected, which were used in all TDA runs. The choice to estimate proper R and G values on the feature set composed of the concatenation of all the considered features (i.e., genetic, environmental, and imaging features) was motivated by the intention to select cover parameters properly capturing the multifactorial features considered in the analysis.

As last step, based on the Euclidean metric of distance and cover parameters, data were partitioned into bins. Density-based spatial clustering (DBSCAN) was performed to cluster subjects in the same bin and tracing an edge between two overlapping clusters (i.e., bins sharing subjects).

#### Graph-based outcome prediction: Step 2

We compared the bootstrapped TDA graphs built on the different feature sets in terms of outcome predictive capability. A quantitative assessment of the associations between the TDA-graph and the three health-related outcome groups was assessed by extracting the SAFE scores. For each outcome group, multiple comparisons using KW tests were performed to test whether the SAFE score distributions were significantly different among the different sets of features over bootstrapped samples.

If significant differences among feature sets were found (p<.05, Bonferroni corrected with n=10, number of feature sets), post-hoc pairwise comparisons were performed to identify possible differences in each pair of feature sets.

From the pairwise comparison, for each outcome, we identified the best predictor as the feature set associated with the highest median value of SAFE score distribution with respect to the other significant pairwise test.

Indeed, the best predictor has the highest median value of SAFE score distribution among pairs and for the highest number of pairs.

#### Feature ranking : Step 3

We ranked features starting from the best predictive feature sets. For each outcome group, Mapper was applied to the best predictive feature set considering the entire sample with the availability of the considered outcomes (n=3052).

On the resulting graph, we developed a new EL metric, which quantifies the variability of a selected variable on all the edges of the graph, by applying a specific function on each pair of nodes connected by an edge. For each pair of nodes connected by an edge, a weighted average of the target variable across the subjects within each node was computed. The subject’s weight was set as inversely proportional to the total number of nodes in which the subject fell.

By considering N as the number of nodes and I as the number of subjects in a node, the node-level (NL) function for the *nth* node was computed as:

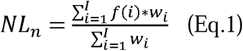

with *f*(*i*) as the target variable value observed in a specific subject, *i* is the selected subject, and *w_i_* is the subject’s weight defined as follows:

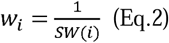

with SW(i) calculated as the number of nodes in which the ith subject is present.

For a pair of nodes (A, B) connected by an edge, the EL metric is calculated as the difference between the corresponding NL metrics:

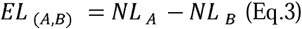

The EL scores (Eq.3) were extracted for each predictive feature and then organized in a matrix of dimensions K x M, where K is the number of graph edges and M is the number of features. Similarly, the EL scores (Eq.3) were extracted for the outcome variables and organized in a vector of dimension K x 1.

To rank the relevance of the predicting features, we fitted linear regression models using the PCs of the ELs of predictors (independent variables) and the ELs of the outcomes of interest (dependent variables). In detail, a PCA was applied to the EL score matrix of the best predictor/best predictive feature set, capturing the edge-related variation in all the best predicting features, previously extracted from the SAFE score graph-outcome prediction analysis. Within this step, the variance of the best predictive features’ EL score matrix was decomposed into the first three PCs representing the directions of the highest variance in the data.

Each PC was separately fed into a linear regression model as an independent variable with the aim to predict the edge related variation of the outcome, identified as the response variable. Inference on the effects of the feature-based PCs on the outcome EL scores were made via t-statistics on the relative beta coefficients. For each outcome, features of the best predictor/best predictive feature set were ranked by the magnitude (from the largest to the smallest) of their PC coefficients extracted from the best predictive PC in the linear regression model. Thus, the most important features of the best predictor were extracted.

## Acknowledgments

This research has been conducted using the UK Biobank Resource under Application Number 56514 “Stratification of health outcomes in mood disorders”. The study was supported by the Italian Ministry of Health (DEPTYPE project, grant n. GR-2019-12370616). PB was partially supported by grants from the Italian Ministry of University and Research (Dipartimenti di Eccellenza Program 2023–2027 – Dept of Pathophysiology and Transplantation, University of Milan), the Italian Ministry of Health (Hub Life Science-Diagnostica Avanzata, HLS-DA, PNC-E3-<u>2022-23683266</u>– CUP: C43C22001630001 / MI-0117; Ricerca Corrente 2024; RF-<u>2019-12371349</u>) and by the Fondazione Cariplo (Made In Family, grant number 2019–3416). EM was partly supported by the Italian Ministry of University and Research (PRIN 2022 PNRR, grant n. P20229MFRC).

## Author contributions

**ET**: Conceptualization, Methodology, Formal analysis, Software, Data curation, Writing— original draft, Writing—review & editing; **AP**: Writing—original draft, Writing—review & editing; **NT**: Methodology, Writing—review & editing; **FC:** Data curation, Methodology, Writing—review & editing**; LF:** Data curation, Methodology, Writing—review & editing**; AMB**: Methodology, Writing—review & editing; **FB**: Writing—review & editing; **CF:** Conceptualization, Project administration, Supervision, Data curation, Methodology, Writing—review & editing, Funding acquisition; **BV:** Conceptualization, Project administration, Supervision, Data curation, Methodology, Writing—review & editing, Funding acquisition; **PB**: Supervision, Writing—review & editing; **EM**: Conceptualization, Project administration; Supervision; Methodology; Writing—original draft, Writing— review & editing, Funding acquisition.

## Data availability statement

The genetic, environmental, brain imaging and clinical outcomes’ datasets generated by UK Biobank analysed during the current study are available via the UK Biobank data access process (see http://www.ukbiobank.ac.uk/register-apply/).

## Code availability statement

The central code of the main work is available upon request at https://osf.io/2se7r/?view_only=9cc533ad4c05449dabfd8613ad9dfdf7.

## References

1. Ferrari, A. J. et al. Burden of Depressive Disorders by Country, Sex, Age, and Year: Findings from the Global Burden of Disease Study 2010. PLoS Med 10, e1001547 (2013).

2. Depression, WHO. Other common mental disorders: global health estimates. Geneva: World Health Organization 24, (2017).

3. Gutiérrez-Rojas, L., Porras-Segovia, A., Dunne, H., Andrade-González, N. & Cervilla, J. A. Prevalence and correlates of major depressive disorder: a systematic review. Braz. J. Psychiatry 42, 657–672 (2020).

4. Chahal, R., Gotlib, I. H. & Guyer, A. E. Research Review: Brain network connectivity and the heterogeneity of depression in adolescence - a precision mental health perspective. J Child Psychol Psychiatry 61, 1282–1298 (2020).

5. Buch, A. M. & Liston, C. Dissecting diagnostic heterogeneity in depression by integrating neuroimaging and genetics. Neuropsychopharmacol. 46, 156–175 (2021).

6. Jentsch, M. C. et al. Biomarker approaches in major depressive disorder evaluated in the context of current hypotheses. Biomark Med 9, 277–297 (2015).

7. Paul, R. et al. Treatment response classes in major depressive disorder identified by model-based clustering and validated by clinical prediction models. Transl Psychiatry 9, 187 (2019).

8. Otte, C. et al. Major depressive disorder. Nat Rev Dis Primers 2, 16065 (2016).

9. Kautzky, A. et al. Combining machine learning algorithms for prediction of antidepressant treatment response. Acta Psychiatr. Scand. 143, 36–49 (2021).

10. Akil, H. et al. Treatment resistant depression: A multi-scale, systems biology approach. Neuroscience & Biobehavioral Reviews 84, 272–288 (2018).

11. Pigoni, A. et al. Can Machine Learning help us in dealing with treatment resistant depression? A review. J Affect Disord 259, 21–26 (2019).

12. Maggioni, E. et al. Common and distinct structural features of schizophrenia and bipolar disorder: The European Network on Psychosis, Affective disorders and Cognitive Trajectory (ENPACT) study. PLoS ONE 12, e0188000 (2017).

13. Bycroft, C. et al. The UK Biobank resource with deep phenotyping and genomic data. Nature 562, 203–209 (2018).

14. Antonucci, L. A. et al. Using combined environmental–clinical classification models to predict role functioning outcome in clinical high-risk states for psychosis and recent-onset depression. Br J Psychiatry 220, 229–245 (2022).

15. Cotrena, C., Damiani Branco, L., Ponsoni, A., Milman Shansis, F. & Paz Fonseca, R. Neuropsychological Clustering in Bipolar and Major Depressive Disorder. J Int Neuropsychol Soc 23, 584–593 (2017).

16. Beijers, L., Wardenaar, K. J., van Loo, H. M. & Schoevers, R. A. Data-driven biological subtypes of depression: systematic review of biological approaches to depression subtyping. Mol Psychiatry 24, 888–900 (2019).

17. Wang, Y. et al. Data-driven clustering differentiates subtypes of major depressive disorder with distinct brain connectivity and symptom features. Br J Psychiatry 219, 606–613 (2021).

18. Chen, X., Dai, Z. & Lin, Y. Biotypes of major depressive disorder identified by a multiview clustering framework. J Affect Disord 329, 257–272 (2023).

19. Kyeong, S., Kim, J.-J. & Kim, E. Novel subgroups of attention-deficit/hyperactivity disorder identified by topological data analysis and their functional network modular organizations. PLoS ONE 12, e0182603 (2017).

20. Romano, D. et al. Topological methods reveal high and low functioning neuro-phenotypes within fragile X syndrome. Human Brain Mapping 35, 4904–4915 (2014).

21. Li, L. et al. Identification of type 2 diabetes subgroups through topological analysis of patient similarity. Sci Transl Med 7, 311ra174 (2015).

22. Yamanashi, T. et al. Topological data analysis (TDA) enhances bispectral EEG (BSEEG) algorithm for detection of delirium. Sci Rep 11, 304 (2021).

23. Nielson, J. L. et al. Topological data analysis for discovery in preclinical spinal cord injury and traumatic brain injury. Nat Commun 6, 8581 (2015).

24. Hyekyoung Lee, Hyejin Kang, Chungo, M. K., Bung-Nyun Kim, & Dong Soo Lee. Persistent Brain Network Homology From the Perspective of Dendrogram. IEEE Trans. Med. Imaging 31, 2267–2277 (2012).

25. Yao, Y. et al. Topological methods for exploring low-density states in biomolecular folding pathways. The Journal of Chemical Physics 130, 144115 (2009).

26. Skaf, Y. & Laubenbacher, R. Topological data analysis in biomedicine: A review. Journal of Biomedical Informatics 130, 104082 (2022).

27. Baryshnikova, A. Systematic Functional Annotation and Visualization of Biological Networks. Cell Systems 2, 412–421 (2016).

28. Liao, T., Wei, Y., Luo, M., Zhao, G.-P. & Zhou, H. tmap: an integrative framework based on topological data analysis for population-scale microbiome stratification and association studies. Genome Biol 20, 293 (2019).

29. Casaclang-Verzosa, G. et al. Network Tomography for Understanding Phenotypic Presentations in Aortic Stenosis. JACC: Cardiovascular Imaging 12, 236–248 (2019).

30. Mullins, N. et al. Polygenic interactions with environmental adversity in the aetiology of major depressive disorder. Psychol. Med. 46, 759–770 (2016).

31. Frodl, T. et al. Effect of hippocampal and amygdala volumes on clinical outcomes in major depression: a 3-year prospective magnetic resonance imaging study. J Psychiatry Neurosci 33, 423–430 (2008).

32. Liston, C., McEwen, B. S. & Casey, B. J. Psychosocial stress reversibly disrupts prefrontal processing and attentional control. Proc Natl Acad Sci U S A 106, 912–917 (2009).

33. Joyce, D. W., Kormilitzin, A., Smith, K. A. & Cipriani, A. Explainable artificial intelligence for mental health through transparency and interpretability for understandability. npj Digit. Med. 6, 6 (2023).

34. Koshiyama, D. et al. White matter microstructural alterations across four major psychiatric disorders: mega-analysis study in 2937 individuals. Mol Psychiatry 25, 883–895 (2020).

35. Zhang, X. et al. Association of Visual Health With Depressive Symptoms and Brain Imaging Phenotypes Among Middle-Aged and Older Adults. JAMA Netw Open 5, e2235017 (2022).

36. Geng, H. et al. Disrupted Structural and Functional Connectivity in Prefrontal-Hippocampus Circuitry in First-Episode Medication-Naïve Adolescent Depression. PLoS ONE 11, e0148345 (2016).

37. Li, W. et al. Effects of the coexistence of late-life depression and mild cognitive impairment on white matter microstructure. Journal of the Neurological Sciences 338, 46–56 (2014).

38. Shim, J.-M., Cho, S.-E., Kang, C.-K. & Kang, S.-G. Low myelin-related values in the fornix and thalamus of 7 Tesla MRI of major depressive disorder patients. Front Mol Neurosci 16, 1214738 (2023).

39. Gianaros, P. J. et al. Potential neural embedding of parental social standing. Soc Cogn Affect Neurosci 3, 91–96 (2008).

40. Dannlowski, U. et al. Childhood maltreatment is associated with an automatic negative emotion processing bias in the amygdala. Hum Brain Mapp 34, 2899–2909 (2013).

41. Ridout, K. K., Ridout, S. J., Price, L. H., Sen, S. & Tyrka, A. R. Depression and telomere length: A meta-analysis. Journal of Affective Disorders 191, 237–247 (2016).

42. Bazaz, M. R., Balasubramanian, R., Monroy-Jaramillo, N. & Dandekar, M. P. Linking the Triad of Telomere Length, Inflammation, and Gut Dysbiosis in the Manifestation of Depression. ACS Chem. Neurosci. 12, 3516–3526 (2021).

43. Price, L. H., Kao, H.-T., Burgers, D. E., Carpenter, L. L. & Tyrka, A. R. Telomeres and Early-Life Stress: An Overview. Biological Psychiatry 73, 15–23 (2013).

44. Xu, L. & Blackburn, E. H. Human Cancer Cells Harbor T-Stumps, a Distinct Class of Extremely Short Telomeres. Molecular Cell 28, 315–327 (2007).

45. Wolkowitz, O. M., Epel, E. S., Reus, V. I. & Mellon, S. H. Depression gets old fast: do stress and depression accelerate cell aging? Depress. Anxiety 27, 327–338 (2010).

46. Mendes-Silva, A. P. et al. Telomere shortening in late-life depression: A potential marker of depression severity. Brain and Behavior 11, e2255 (2021).

47. Hartmann, N., Boehner, M., Groenen, F. & Kalb, R. Telomere length of patients with major depression is shortened but independent from therapy and severity of the disease. Depress. Anxiety 27, 1111–1116 (2010).

48. Berk, M. et al. Comorbidity between major depressive disorder and physical diseases: a comprehensive review of epidemiology, mechanisms and management. World Psychiatry 22, 366–387 (2023).

49. Hare, D. L., Toukhsati, S. R., Johansson, P. & Jaarsma, T. Depression and cardiovascular disease: a clinical review. European Heart Journal 35, 1365–1372 (2014).

50. Grippo, A. J. & Johnson, A. K. Stress, depression and cardiovascular dysregulation: A review of neurobiological mechanisms and the integration of research from preclinical disease models: Review. Stress 12, 1–21 (2009).

51. Penninx, B. W. J. H. Depression and cardiovascular disease: Epidemiological evidence on their linking mechanisms. Neuroscience & Biobehavioral Reviews 74, 277–286 (2017).

52. Friedman, J. I. et al. Brain imaging changes associated with risk factors for cardiovascular and cerebrovascular disease in asymptomatic patients. JACC Cardiovasc Imaging 7, 1039–1053 (2014).

53. Sheline, Y. I. 3D MRI studies of neuroanatomic changes in unipolar major depression: the role of stress and medical comorbidity. Biological Psychiatry 48, 791–800 (2000).

54. Gold, S. M. et al. Comorbid depression in medical diseases. Nat Rev Dis Primers 6, 69 (2020).

55. Valenza, G. Depression as a cardiovascular disorder: central-autonomic network, brain-heart axis, and vagal perspectives of low mood. Front Netw Physiol 3, 1125495 (2023).

56. Tonhajzerova, I. et al. Major depressive disorder at adolescent age is associated with impaired cardiovascular autonomic regulation and vasculature functioning. International Journal of Psychophysiology 181, 14–22 (2022).

57. Mayberg, H. S. et al. Cingulate function in depression: a potential predictor of treatment response. NeuroReport 8, 1057–1061 (1997).

58. Fonseka, T. M., MacQueen, G. M. & Kennedy, S. H. Neuroimaging biomarkers as predictors of treatment outcome in Major Depressive Disorder. Journal of Affective Disorders 233, 21–35 (2018).

59. Klok, M. P. C., Van Eijndhoven, Philip. F., Argyelan, M., Schene, A. H. & Tendolkar, I. Structural brain characteristics in treatment-resistant depression: review of magnetic resonance imaging studies. BJPsych open 5, e76 (2019).

60. Kotoula, V., Evans, J. W., Punturieri, C., Johnson, S. C. & Zarate, C. A. Functional MRI markers for treatment-resistant depression: Insights and challenges. in Progress in Brain Research vol. 278 117–148 (Elsevier, 2023).

61. Ferri, J. et al. Blunted amygdala activity is associated with depression severity in treatment-resistant depression. Cogn Affect Behav Neurosci 17, 1221–1231 (2017).

62. Vai, B. et al. Fronto-limbic effective connectivity as possible predictor of antidepressant response to SSRI administration. European Neuropsychopharmacology 26, 2000–2010 (2016).

63. Fabbri, C. et al. Genetic and clinical characteristics of treatment-resistant depression using primary care records in two UK cohorts. Mol Psychiatry 26, 3363–3373 (2021).

64. Fry, A. et al. Comparison of Sociodemographic and Health-Related Characteristics of UK Biobank Participants With Those of the General Population. American Journal of Epidemiology 186, 1026–1034 (2017).

## Methods References

65. Davis, K. A. S. et al. Mental health in UK Biobank – development, implementation and results from an online questionnaire completed by 157 366 participants: a reanalysis. BJPsych open 6, e18 (2020).

66. Smith, D. J. et al. Prevalence and Characteristics of Probable Major Depression and Bipolar Disorder within UK Biobank: Cross-Sectional Study of 172,751 Participants. PLoS ONE 8, e75362 (2013).

67. Alfaro-Almagro, F. et al. Image processing and Quality Control for the first 10,000 brain imaging datasets from UK Biobank. NeuroImage 166, 400–424 (2018).

68. Hariri, A. R., Tessitore, A., Mattay, V. S., Fera, F. & Weinberger, D. R. The Amygdala Response to Emotional Stimuli: A Comparison of Faces and Scenes. NeuroImage 17, 317–323 (2002).

69. Jiang, R. et al. A functional connectome signature of blood pressure in >30 000 participants from the UK biobank. Cardiovascular Research 119, 1427–1440 (2023).

70. Codd, V. et al. Measurement and initial characterization of leukocyte telomere length in 474,074 participants in UK Biobank. Nat Aging 2, 170–179 (2022).

71. Ge, T., Chen, C.-Y., Ni, Y., Feng, Y.-C. A. & Smoller, J. W. Polygenic prediction via Bayesian regression and continuous shrinkage priors. Nat Commun 10, 1776 (2019).

72. Chang, C. C. et al. Second-generation PLINK: rising to the challenge of larger and richer datasets. GigaSci 4, 7 (2015).

73. Milaneschi, Y., Lamers, F., Berk, M. & Penninx, B. W. J. H. Depression Heterogeneity and Its Biological Underpinnings: Toward Immunometabolic Depression. Biological Psychiatry 88, 369–380 (2020).

74. Fabbri, C., Mutz, J., Lewis, C. M. & Serretti, A. Depressive symptoms and neuroticism-related traits are the main factors associated with wellbeing independent of the history of lifetime depression in the UK Biobank. Psychol. Med. 53, 3000–3008 (2023).

75. Brailean, A., Curtis, J., Davis, K., Dregan, A. & Hotopf, M. Characteristics, comorbidities, and correlates of atypical depression: evidence from the UK Biobank Mental Health Survey. Psychol. Med. 50, 1129–1138 (2020).

76. Wills, P. & Meyer, F. G. Metrics for graph comparison: A practitioner’s guide. PLoS ONE 15, e0228728 (2020).

