## Supplementary material for "Clinical stratification of Major Depressive Disorder in the UK Biobank: A gene-environment-brain Topological Data Analysis": https://osf.io/2se7r/?view_only=9cc533ad4c05449dabfd8613ad9dfdf7

### ***Supplementary Information***

Table of contents:

Supplementary Methods……………………………………………………………………………
Supplementary Figure S1…………………………………………………………………………..
Supplementary Figure S2…………………………………………………………………………..
Supplementary Table S1……………………………………………………………………………
Supplementary Table S2……………………………………………………………………………
Supplementary Table S3……………………………………………………………………………
Supplementary Table S4……………………………………………………………………………

### Supplementary Methods

**Imaging measures pre-processing steps.**

The processing and quality control (QC) pipelines applied to extracted brain imaging predictors were previously described by [1]. Briefly, part of the MRI-derived features extraction's pipelines and related parameters were reported as follow.

***sMRI***. T1-weighted imaging was performed by using a three-dimensional magnetization-prepared rapid acquisition with gradient echo sequence (MPRAGE) (resolution: 1.0 x 1.0 x 1.0 mm; field-of-view matrix: 208 x 256 x 256; inversion time, 880 msec; repetition time, 2000 msec). T1-weighted data were first segmented by using FAST (version 4.1; Automated Segmentation Tool in FMRIB) to extract gray matter, white matter, and cerebrospinal fluid. Subcortical structures were extracted by using FIRST (version 5.0; Integrated Registration and Segmentation Tool in FMRIB). Cortical regional volumes, cortical thickness and surface area obtained using Freesurfer according on Desikan-Killiany atlas were employed (UKB Data fields within the Category: 1101, Regional grey matter volumes (FAST); 1102, Subcortical volumes (FIRST); 196, Freesurfer DKT).

***dMRI***. Diffusion tensor imaging meaures (DTI) imaging was performed using echo-planar, single-shot Stejskal-Tanner pulse sequence (echo time msec, 92) to obtain 36 sections (resolution, 2.0 x 2.0 x 2.0 mm; field-of-view matrix: 104 x 104 x 72 matrix) in 50 distinct diffusion-weighted directions (*b* values, 1000 and 2000 sec/ mm2). DTI in individual white matter tracts were extracted using FMRIB Software Library (FSL) tools (http://www.fmrib.ox.ac.uk/fsl). DTI variables extracted includes fractional anisotropy (FA) and mean diffusivity (MD) as well as orientation dispersion index (ODI), measures such as Intracellular Volume Fraction (ICVF), the Isotropic Compartment Volume Fraction (ISOVF), axial diffusivity (AD), radial diffusivity (RD) and L1 direction. All DTI measures were averaged within 48 distinct tract ROIs defined using the Johns Hopkins University tract atlas (UKB Data fields within the Category 134, dMRI skeleton).

***rs-fMRI***. rs-fMRI was performed using gradient echo echo-planar imaging (GE EPI) (resolution: 2.4 x 2.4 x 2.4 mm; field-of-view matrix: 88 x 88 x 64; duration time, 6 min; repetition time, 0.735 sec; echo time, 39 msec; flip angle, 52°). rs-fMRI network matrices were employed, extracted from the imaging-derived phenotypes that were processed by the UK Biobank imaging project team. Data preprocessing, group independent component analysis (ICA) parcellation, and connectivity estimation were carried out using FSL packages (http://biobank.ctsu.ox.ac.uk/ crystal/refer.cgi?id=1977) by the UK Biobank. Network matrices obtained using group-ICA with dimensionality of D=100 independent component (ICs) and non-artificial components were considered. This identified 55 components in 100-D that remained for further analysis. Connections between pairs of ICs were extracted by considering partial correlation as measure of functional connections among components obtaining a 55x55 partial correlation matrix for each individual. From the matrix of connectivity estimates extracted at subject-level with dimension 55 ICs x 55 ICs were extracted the parameters of strength of positive (Spos) and negative weights (Sneg) for each network "node", resulting in 55 Spos and 55 Sneg measures. Hence, each of the 55 ICs were visualized based on the UkBiobank's online visualization tool (https://www.fmrib.ox.ac.uk/datasets/ukbiobank/group_means/rfMRI_ICA_d100.html) and grouped into 9 typical large functional networks, in accordance with previous studies[2] (UKB Data fields within the Category 203, Resting functional brain MRI).

***t-fMRI***. t-fMRI was performed using GE EPI (resolution: 2.4 x 2.4 x 2.4 mm; field-of-view matrix: 88 x 88 x 64; duration time, 4 min; repetition time, 0.735 sec; echo time, 39 msec; flip angle, 52°). Task-based activation during implicit emotion processing of the Hariri faces/shapes "emotion" task [3] were extracted. Specifically, the participants were presented with blocks of trials and asked to decide either which of two faces presented on the bottom of the screen match the face at the top of the screen, or which of two shapes presented at the bottom of the screen match the shape at the top of the screen. The faces have either angry or fearful expressions. These variables include Median blood-oxygen-level-dependent (BOLD) effect and 90$^{th}$ percentile (in group-defined amygdala activation mask) for faces-shapes contrast, Median BOLD effect and 90$^{th}$ percentile (in group-defined mask) for faces activation, Median BOLD effect and 90$^{th}$ percentile (in group-defined mask) for faces-shapes contrast, Median BOLD effect and 90$^{th}$ percentile (in group-defined mask) for shapes activation (UKB Data fields within the Category 106, Task functional brain MRI).

**Genotyping, quality control and imputation**

Genome-wide genotyping was performed all Uk Biobank was performed using two highly overlapping arrays covering ~600.000 markers. Autosomal genotype data underwent centralized quality control to adjust for possible array effects, batch effects, plate effects, and departures from Hardy-Weinberg equilibrium (HWE)[4]. SNPs were further excluded based on missingness (> 0.05) and on Hardy Weinberg equilibrium (p <10-8). Individuals were removed for high levels of missingness (> 0.05) or abnormal heterozygosity (as defined during centralised quality control), relatedness of up to third-degree kinship[5] (KING r < 0.044) or phenotypic and genotypic gender discordance. Population structure within the UK Biobank cohort was assessed using principal component analysis, with European ancestry defined by 4-means clustering on the first two genetic principal components[6]. A two-stage imputation was performed using the Haplotype Reference Consortium (HRC) and UK10K reference panels[4,7,8]. Poor imputed variants were excluded[7] (INFO ≤ 0.4).

#### Supplementary Figures

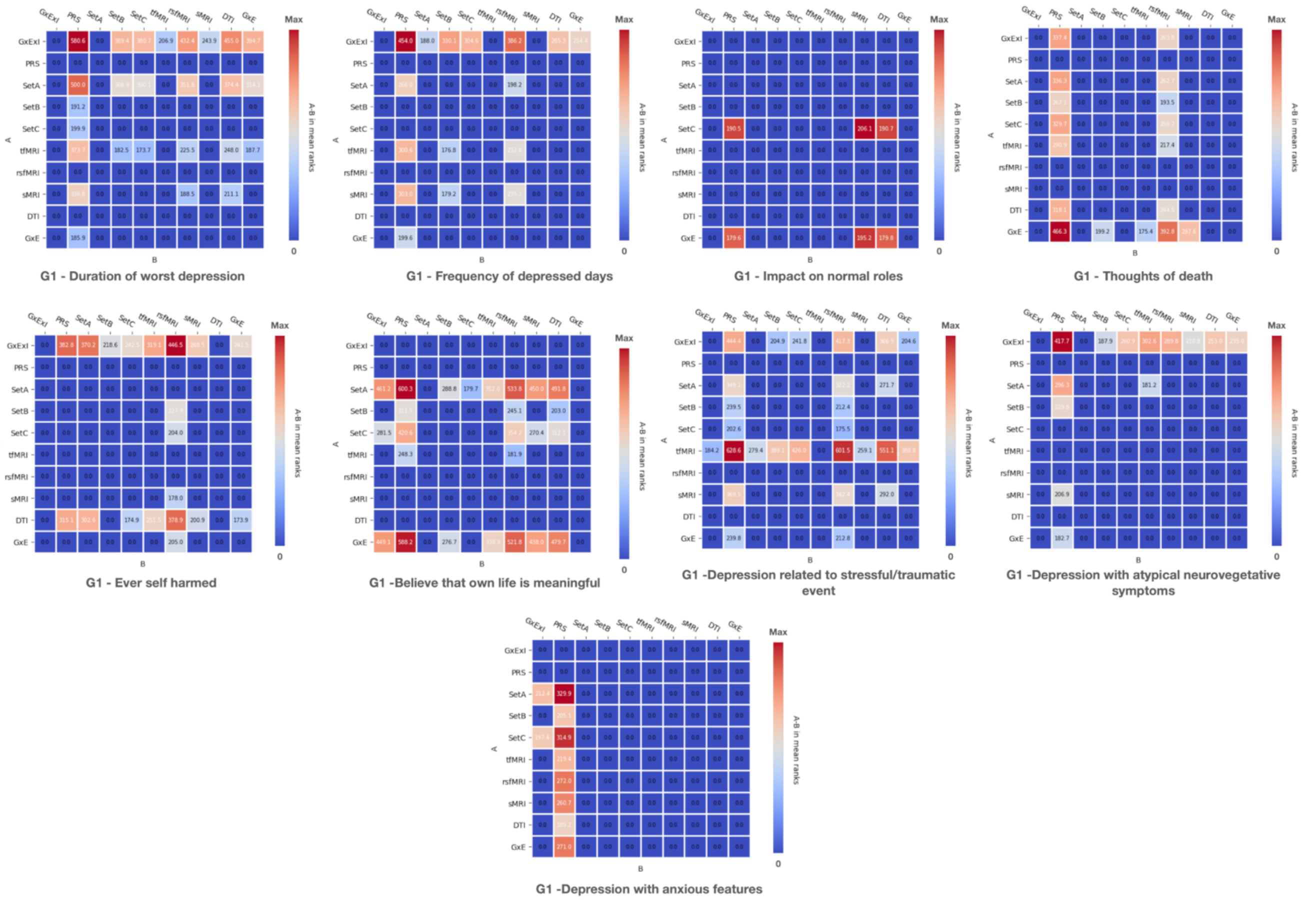


**Figure S1,** Heatmaps of G1 outcomes representative of post-hoc pairwise comparison of SAFE score distribution among feature sets. Each matrix is characterized by rows and columns equals to the feature sets considered (10 in total). Each matrix serves for representing the post-hoc comparison of the SAFE score distribution's results between feature sets (A: feature sets on the Y axis, B: feature sets on the X axis). Each entry represents the "A-B mean ranks" that stands for the difference in the median value of SAFE score distribution of a pair of feature sets, where A indicate the first feature set in the difference and B the second feature in the difference.

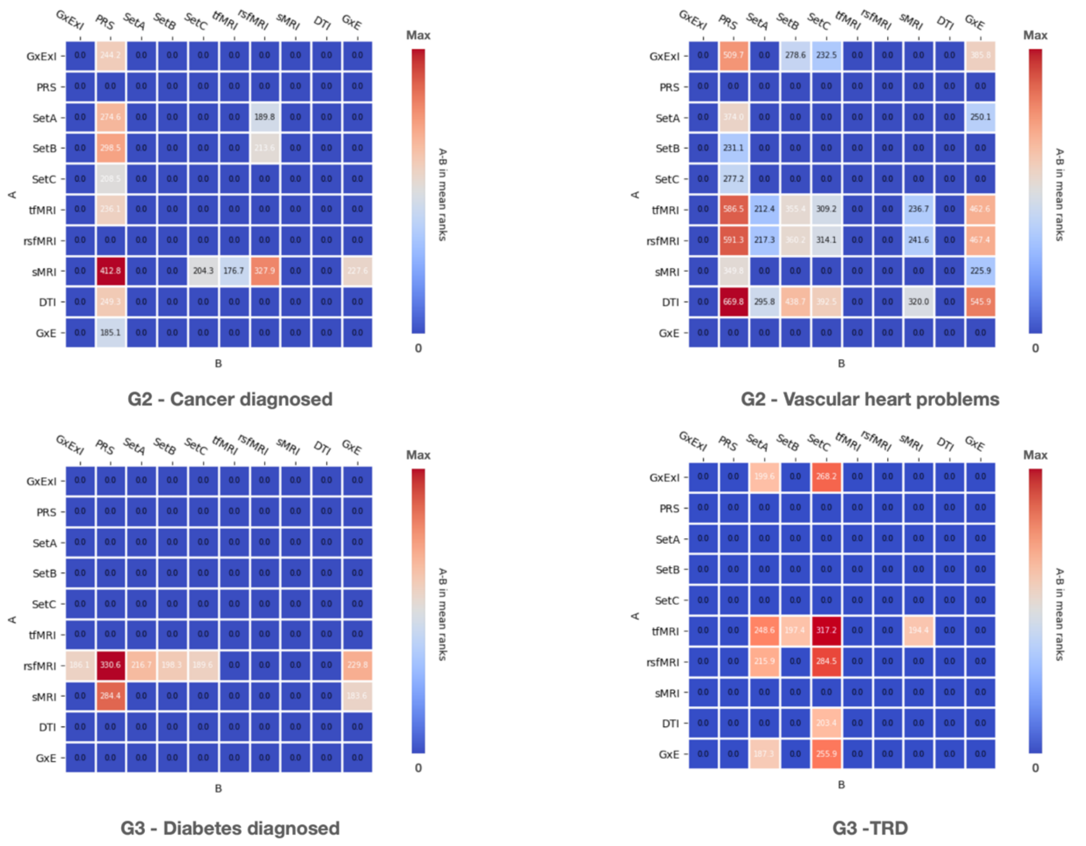


**Figure S2**, Heatmaps of G2 and G3 outcomes representative of post-hoc pairwise comparison of SAFE score distribution among feature sets. Each matrix is characterized by rows and columns equals to the feature sets considered (10 in total). Each matrix serves for representing the post-hoc comparison of the SAFE score distribution's results between feature sets (A: feature sets on the Y axis, B: feature sets on the X axis). Each entry represents the "A-B mean ranks" that stands for the difference in the median value of SAFE score distribution of a pair of feature sets, where A indicate the first feature set in the difference and B the second feature in the difference

#### Supplementary Tables

**Table S1.** Resting-state networks associated with the 55 non-artefactual ICs generated from group-ICA of UKB cohort.

| **RSNs** | Non-artefactual ICs included |
|---|---|
| VAN | IC-2, -15, -37, -45, -47 |
| VIS | IC-1, -3, -16, -8, -14 |
| DMNp | IC-5, -10, -11, -19, -36 |
| SMN | IC-6, -20, -22, -30, -32, -35 |
| DMNa | IC-7, -13, -21, -28, -52, -55 |
| Sub&Cereb | IC-38, -17, -23, -53, -54 |
| DAN | IC-5, -18, -27, -34, -39, -40, -41, -42 |
| TempPar | IC-9, -33, -43, -48, -50, -51 |
| FPN | IC-12, -24, -25, -26, -29, -31, -44, -46, -49 |

Abbreviations: RSNs, resting-state networks; ICs, independent components; DMNa, default mode network anterior; DMNp, default mode network posterior; SMN, somatomotor network; Sub&Cereb, subcortical and cerebellar network; TempPar, temporoparietal network; VAN, ventral attention network; DAN, dorsal attention network; VIS, visual network ; FPN, frontoparietal network.

**Table S2.** Genome-wide association studies used to estimate the effect of genetic variants associated with various traits and further for the calculation of polygenic risk scores.

| **Trait** | **Type of trait** | **N cases** | **N controls** | **Ethnicity** | **PMID** |
|---|---|---|---|---|---|
| MDD | Binary | 45,591 | 97,674 | European | 29700475 |
| BD | Binary | 20,352 | 31,358 | European | 31043756 |
| Schizophrenia | Binary | 40,675 | 64,643 | European | 29483656 |
| ADHD | Binary | 20,183 | 35,191 | European | 30478444 |
| Anorexia nervosa | Binary | 16,992 | 55,525 | European | 31308545 |
| Autism | Binary | 18,381 | 27,969 | European | 30804558 |
| Alcohol Dependence | Binary | 14,904 | 37,944 | Mixed | 30482948 |
| Daily alcohol consumption | Continuous | 70,460 | | European | 27911795 |
| BMI | Continuous | 322,154 | | European | 25673413 |
| Coronary artery disease | Binary | 60,801 | 123,504 | European e Asiatic | 26343387 |
| C reactive protein | Binary | 204,402 | | European | 30388399 |

| | | | | | |
|---|---|---|---|---|---|
| Type 2 diabetes mellitus | Binary | 26,676 | 132,532 | Mixed | 28566273 |
| Glycated hemoglobin | Continuous | 123,665 | | European | 28898252 |
| N sigarette / day | Continuous | 38,181 | | European | 20418890 |
| Triglycerides | Continuous | 1,654,960 | | Mixed | 34887591 |
| LDL cholesterol | Continuous | 1,654,960 | | Mixed | 34887591 |
| HDL cholesterol | Continuous | 1,654,960 | | Mixed | 34887591 |
| Years of education | Continuous | 766,345 | | European | 30038396 |

Abbreviations: Bipolar disorder (BD-PRS), schizophrenia (SCZ-PRS), MDD (DEPR07-PRS), anorexia nervosa (ANOR07-PRS), autism (AUTI07-PRS), attention deficit hyperactivity disorder (ADHD05-PRS). PRS related to the following non-psychiatric traits were extracted: smoking (SMOK03-PRS), alcohol dependence (ALCD01_PRS), C reactive protein (CRP-PRS), years of education (education-PRS), glycated hemoglobin (hb1ac-PRS), triglycerides (TRGL_PRS), LDL cholesterol (LDL-PRS), coronary artery disease (CAD-PRS), alcohol consumption (ALCO01_PRS), body mass index (BMI-PRS), HDL cholesterol (HDL_PRS), type 2 diabetes mellitus (DM-PRS), neuroticism (Neuroticism-PRS).

**Table S3.** Environmental characteristics included in SetA and relative UKB code.

| **Environmental Set A** | | | |
|---|---|---|---|
| Name | UKB Data field or definition | Variable Domain original | Variable Domain modified |
| Frequency of friend / family visits | 1031 | 7 levels: 1 almost every day, 2 2-4 times a week, 3 about once a week, 4 about once a month, 5 once every few months, 6 never or almost never, 7 no family/friends outside household | Binary: 0 no frequent visit (including once every few months, never or almost never, no family/friends outside household), 1 frequent visit (including almost daily, 2-4 times a week, about once a week, about once a month). |
| Leisure / social activities | 6160 | 5 levels: 1 Sports clubs or gym, 2 pub or social club, 3 religious group, 4 adult education class, 5 other group of activity | Binary: 0 never, 1 any type of leasure/social activities (including: sports clubs or gym, pub or social club, religious group, adult education class, other group of activity) |
| Able to confide | 2110 | 6 levels: 5 almost every day, 4 2.4 times a week, 3 about once a week, 2 about once a month, 1 once every few months, 0 never or almost never. | Binary: 0 lack of social support (including: 2.4 times a week, about once a week, about once a month, once every few months, never or almost never), 1 regular social support ( almost every day) |

| | | | |
|---|---|---|---|
| Morning/evening person (chronotype) | 1180 | 4 levels: 1 definitely a “morning person”, 2 more a “morning person” then “evening person”, 3 more an “evening person” than a “morning person”, 4 definitely an “evening person”. | Binary: 0 morning person (including: more an “evening person” than a “morning person”, definitely an “evening person”), 1 evening person (including: definitely a “morning person”, more a “morning person” then “evening person”) |
| Summed MET minutes per week for all activity | 22040 | Continuous | / |
| Z adjusted T/S log (i.e., Telomeres length) | 22192 | Continuous | / |
| Neuroticism-EPQ RS | Derived from 10.1038/mp.2016.49 | / | 12 levels extracted from 12 items of neuroticism scale from the Eysenck Personality Questionnaire-Revised Short Form. |

**Abbreviations:** MET, metabolic equivalent task.

**Table S4.** Environmental characteristics included in SetB and relative UKB code.

| **Environmental Set B** | | | |
|---|---|---|---|
| Name | UKB Data field or definition | Variable Domain original | Variable Domain modified |
| Felt love as a child | 20489 | 5 levels: 0 never, 1 rarely true, 2 sometimes true, 3 often, 4 very often | / |
| Physically abused by family as a child | 20488 | 5 levels: 0 never, 1 rarely true, 2 sometimes true, 3 often, 4 very often | / |
| Felt hated by family member as a child | 20487 | 5 levels: 0 never, 1 rarely true, 2 sometimes true, 3 often, 4 very often | / |
| Sexually molested as a child | 20490 | 5 levels: 0 never, 1 rarely true, 2 sometimes true, 3 often, 4 very often | / |
| Someone to take to doctor when needed as a child | 20491 | 5 levels: 0 never, 1 rarely true, 2 sometimes true, 3 often, 4 very often | / |
| Been in a confiding relationship as an adult | 20522 | 5 levels: 0 never, 1 rarely true, 2 sometimes true, 3 often, 4 very often | / |
| Physical violence by partner or ex-partner as an adult | 20523 | 5 levels: 0 never, 1 rarely true, 2 sometimes true, 3 often, 4 very often | / |
| Belittlement by partner or ex-partner as an adult | 20521 | 5 levels: 0 never, 1 rarely true, 2 sometimes true, 3 often, 4 very often | / |
| Sexual interference by partner or ex-partner without consent as an adult | 20524 | 5 levels: 0 never, 1 rarely true, 2 sometimes true, 3 often, 4 very often | / |
| Able to pay rent/mortgage as an adult | 20525 | 5 levels: 0 never, 1 rarely true, 2 sometimes true, 3 often, 4 very often | / |
| Victim of sexual assault | 20531 | 3 levels: 0 never, 1 yes but not in the last 12 months, 2 yes within the last 12 months. | Binary: 0 never, 1 it happened (including: yes but not in the last 12 months, yes within the last 12 months) |

| | | | |
|---|---|---|---|
| Victim of physically violent crime | 20529 | 3 levels: 0 never, 1 yes but not in the last 12 months, 2 yes within the last 12 months. | Binary: 0 never, 1 it happened (including: yes but not in the last 12 months, yes within the last 12 months) |
| Been in serious accident believed to be life-threatening | 20526 | 3 levels: 0 never, 1 yes but not in the last 12 months, 2 yes within the last 12 months. | Binary: 0 never, 1 it happened (including: yes but not in the last 12 months, yes within the last 12 months) |
| Witnessed sudden violent death | 20530 | 3 levels: 0 never, 1 yes but not in the last 12 months, 2 yes within the last 12 months. | Binary: 0 never, 1 it happened (including: yes but not in the last 12 months, yes within the last 12 months) |
| Diagnosed with life-threatening illness | 20528 | 3 levels: 0 never, 1 yes but not in the last 12 months, 2 yes within the last 12 months. | Binary: 0 never, 1 it happened (including: yes but not in the last 12 months, yes within the last 12 months) |
| Been involved in combat or exposed to war-zone | 20527 | 3 levels: 0 never, 1 yes but not in the last 12 months, 2 yes within the last 12 months. | Binary: 0 never, 1 it happened (including: yes but not in the last 12 months, yes within the last 12 months) |
| Alcohol intake frequency | 1558 | 6 levels: 1 daily or almost daily, 2 three or four times a week, 3 once or twice a week, 4 once to three times a month, 5 special occasion only, 6 never. | 3 levels: 0 never, 1 special occasion only or sometimes a month, 2 sometimes a week, 3 almost daily |
| Childhood adverse events | Derived from doi: 10.1016/j.ynstr.2022.100447 based on: 20489, 20488, 20487, 20490, 20491 | / | 5 levels. |
| Adulthood stress | Derived from 10.1016/j.ynstr.2022.100447 based on: 20487, 20488, 20489, 20490, 20491 | / | 4 levels. |
| Serious illness, injury, assault to yourself or assault of a close relative in the last 2 years | 6145 | 6 levels: 1 serious illness, injury or assault to yourself, 2 serious illness, injury or assault of a close relative, 3 death of a close relative, 4 death of a spouse or partner, 5 marital separation/divorce, 6 financial difficulties | Binary: 1 if it happened (including: serious illness, injury or assault to yourself, serious illness, injury or assault of a close relative ), 0 None (including all the other categories). |
| Death of a close relative or death of a spouse or partner in the last 2 years | 6145 | 6 levels: 1 serious illness, injury or assault to yourself, 2 serious illness, injury or assault of a close relative, 3 death of a close relative, 4 death of a spouse or partner, 5 marital separation/divorce, 6 financial difficulties | Binary: 1 if it happened (including: death of a close relative, death of a spouse or partner), 0 None ( including all the other categories), |
| Stress related to marital separation/divorce or to financial difficulties in the last 2 years | 6145 | 6 levels: 1 serious illness, injury or assault to yourself, 2 serious illness, injury or assault of a close relative, 3 death of a close relative, 4 death of a spouse or partner, 5 marital separation/divorce, 6 financial difficulties | Binary: 1 if it happened (including: marital separation/divorce, financial difficulties), 0 None ( including all the other categories). |

| | | | |
|---|---|---|---|
| Ever taken cannabis | 20453 | 5 levels: 0 no, 1 yes 1-2 times, 2 yes 3-10 times, 3 yes 11-100 times, 4 yes more than 100 times. | 3 levels: 0 never, 1 if at maximum 10 times, 2 if more than 10 times |
| Smoking status | 20116 | 3 levels: 0 if never, 1 if previous, 2 if current | / |
| Major dietary changes because of illness in the last 5 years | 1538 | 3 levels: 0 no, 1 yes because of illness, 2 yes because of other reasons. | Binary: 0 never, 1 if it happened. |
| Major dietary changes because other reason in the last 5 years | 1538 | 3 levels: 0 no, 1 yes because of illness, 2 yes because of other reasons. | Binary: 0 never, 1 if it happened. |

**Table S5**. Outcomes included in each group with the relative UkBioBank code.

| Group 1 (G1) | | | |
|---|---|---|---|
| Name | UKB Data field or definition | Variable Domain original | Variable Domain modified |
| O1: Duration of worst depression | 20438 | 3 levels: 1=1-2 Below 3 months, 2=3-4 below 1 year=2, 3=5,6 up to one year | |
| O2: Frequency of depressed days during worst episode of depression | 20439 | 3 levels: 1= less often, 2 = almost every day, 3= every day | |
| O3: Impact on normal roles during worst period of depression | 20440 | 3 levels: 0 not at all, 1 a little, 2 somewhat, 3 a lot | |
| O4: Thoughts of death during worst depression | 20437 | Binary: 0 none, 1 if it happened | |
| O5: Ever self harmed | 20480 | Binary: 0 none, 1 if it happened | |
| O6: Belief that owns life is meaningful | 20460 | 5 levels: 1 Not at all, 2 A little, 3 A moderate amount, 4 Very much, 5 An extreme amount | |
| O7: Depression possibly related to stressful or traumatic event | 20447 | Binary: 0 condition not present, 1 condition present | |
| O8: Depression with atypical neurovegetative symptoms | Derived from https://doi.org/10.1038%2Fs41386-021-01059-6. | Binary: 0 condition not present, 1 condition present | |
| O9: Depression with anxious features | Derived from https://doi.org/10.1038%2Fs41386-021-01059-6. | Binary: 0 condition not present, 1 condition present | |
| **Group 2 (G2)** | | | |
| Name | UKB Data field or definition | Variable Domain original | Variable Domain modified |
| O1: Cancer diagnosed by doctor | 2453 | Binary: 0 not diagnosed, 1 diagnosed | / |
| O2: Vascular heart problems | 6150 | 3 levels: 2 if HBP, heart attack, stroke or angina, 1 if HBP, 0 if none | 3 levels: 0=None, 1=HBP, 2=HBP with Stroke, or Angina or Heart Attack |
| O3: Diabetes diagnosed by doctors | 2443 | Binary: 0 not diagnosed, 1 diagnosed | / |
| **Group 3 (G3)** | | | |
| O1: Treatment resistant depression | Derived from 10.1038/s41380-021-01062-9. | / | Binary: 0 condition not present, 1 condition present |

Abbreviations: G, group of clinical outcomes; O, clinical outcome.

**Table S6**. List of socio-demographic characteristics of the sample included with the relative UkBioBank code.

| UKB Data field or definition | Name |
|---|---|
| 21003 | Age at baseline |
| 31 | Sex |
| 21000 | Ethnicity |
| 21001 | BMI |
| 48 | WC |

Abbreviations: BMI, Body mass index; WC, waist circumference.

**Table S7**. List of drugs included in antidepressant medications with the relative UkBioBank code.

| UKB Data field or definition | Name |
|---|---|
| 1140879616 | amitriptyline |
| 1140921600 | citalopram |
| 1140879540 | fluoxetine |
| 1140867878 | sertraline |
| 1140916282 | venlafaxine |
| 1140909806 | dosulepin |
| 1140867888 | paroxetine |
| 1141152732 | mirtazapine |
| 1141180212 | escitalopram |
| 1140879634 | trazodone |
| 1140867876 | prozac |
| 1140882236 | seroxat |
| 1141190158 | cipralex |
| 1141200564 | duloxetine |
| 1140867726 | lofepramine |
| 1140879620 | clomipramine |
| 1140867818 | nortriptyline |
| 1140879630 | imipramine |
| 1140879628 | dothiepin |
| 1141151946 | cipramil |
| 1140867624 | prothiaden |
| 1140867756 | trimipramine |
| 1140867884 | lustral |
| 1141151978 | reboxetine |
| 1141152736 | zispin |
| 1141201834 | cymbalta |
| 1140867690 | anafranil |
| 1140867640 | doxepin |
| 1140867920 | moclobemide |
| 1140867850 | phenelzine |
| 1140879544 | fluvoxamine |
| 1141200570 | yentreve |
| 1140867934 | triptafen |
| 1140867758 | surmontil |
| 1140867914 | tranylcypromine |
| 1140867820 | allegron |
| 1141151982 | edronax |
| 1140882244 | molipaxin |
| 1140879556 | mianserin |
| 1140867852 | nardil |
| 1140867860 | faverin |
| 1140917460 | nefazodone |
| 1140867938 | amitriptyline+chlordiazepoxide |
| 1140867856 | isocarboxazid |
| 1140867922 | manerix |
| 1140910820 | maoi |
| 1140882312 | sinequan |
| 1140867944 | tranylcypromine+trifluoperazine |
| 1140867784 | ludiomil |
| 1140867812 | norval |

| | |
|---|---|
| 1140867668 | tryptizol |
| 1140867940 | fluphenazine hydrochloride+nortriptyline |
| 1140867948 | amitriptyline hydrochloride+perphenazine 10mg/2mg tablet |
| 1140867774 | amoxapine |
| 1141174756 | oxactin 20mg capsule |
| 1140916288 | efexor 37.5mg tablet |
| 1140867628 | prepadine 25mg capsule |
| 1140882310 | gamanil 70mg tablet |
| 1140867784 | ludiomil |
| 1140867930 | motival |
| 1140867712 | tofranil |
| 1140867916 | parnate |

#### References


1. Alfaro-Almagro, F. *et al.* Image processing and Quality Control for the first 10,000 brain imaging datasets from UK Biobank. *NeuroImage* **166**, 400–424 (2018).
2. Jiang, R. *et al.* A functional connectome signature of blood pressure in >30 000 participants from the UK biobank. *Cardiovasc. Res.* **119**, 1427–1440 (2023).
3. Hariri, A. R., Tessitore, A., Mattay, V. S., Fera, F. & Weinberger, D. R. The Amygdala Response to Emotional Stimuli: A Comparison of Faces and Scenes. *NeuroImage* **17**, 317–323 (2002).
4. Bycroft, C. *et al.* The UK Biobank resource with deep phenotyping and genomic data. *Nature* **562**, 203–209 (2018).
5. Manichaikul, A. *et al.* Robust relationship inference in genome-wide association studies. *Bioinformatics* **26**, 2867–2873 (2010).
6. The International Consortium of Blood Pressure (ICBP) 1000G Analyses *et al.* Genome-wide association analysis identifies novel blood pressure loci and offers biological insights into cardiovascular risk. *Nat. Genet.* **49**, 403–415 (2017).
7. the Haplotype Reference Consortium. A reference panel of 64,976 haplotypes for genotype imputation. *Nat. Genet.* **48**, 1279–1283 (2016).
8. The UK10K Consortium *et al.* The UK10K project identifies rare variants in health and disease. *Nature* **526**, 82–90 (2015).
